# Multivariate genome-wide association study dissects shared biology and disorder-specific loci across internalizing spectrum in millions of ancestrally diverse participants

**DOI:** 10.64898/2026.03.27.26348717

**Authors:** Dan Qiu, Zhongzheng Mao, Jun He, Ziang Xu, Chang Liu, David Davtian, Qianyu Chen, Sefayet Karaca, 23andMe Research Team, Brenda Cabrera-Mendoza, Renato Polimanti

## Abstract

The extent of shared and disorder-specific etiology among generalized anxiety disorder (GAD), major depressive disorder (MDD), and posttraumatic stress disorder (PTSD) remains unclear. Leveraging multiple cohorts, we conducted a multivariate and multi-ancestry genome-wide association study of GAD (N=1,358,762), MDD (N=3,601,629), and PTSD (N=1,617,876). We identified 248 loci associated with the latent internalizing disorder factor (INT), 591 with MDD, 237 with PTSD, and 109 with GAD. While GAD and PTSD genetic risk demonstrated strong overlap with the INT factor, 38% of MDD genetic signals were disorder-specific. Cross-population fine-mapping uncovered >450 causal variants, and the subsequent multi-omics characterization linked them to >1,250 genes, including both novel shared and disorder-specific loci. Considering the high-confidence findings converging across analytic approaches, we observed that the genetic liability shared across internalizing spectrum is driven by broadly acting cellular and regulatory mechanisms, whereas disorder-specific genetic risk reflects more specialized perturbations in neurodevelopmental, synaptic, and stress-responsive pathways.

## MAIN

Generalized anxiety disorder (GAD), major depressive disorder (MDD), and posttraumatic stress disorder (PTSD) represent some of the most prevalent and disabling psychiatric conditions worldwide.^1^ Although these disorders are clinically distinct, they frequently co-occur, share overlapping symptom dimensions, and show substantial genetic correlations, suggesting the presence of a shared biological architecture.^2^ Over the past decade, large-scale genome-wide association studies (GWAS) have identified many loci associated with each disorder, greatly expanding our understanding of the polygenic basis of the internalizing spectrum.^3–6^

Recently, a large-scale cross-disorder GWAS study conducted by the Psychiatric Genomics Consortium (PGC) provided a comprehensive view of the genetic landscape across 14 psychiatric disorders^2^. This analysis of more than one million cases identified latent genomic factors that explain genetic variance shared across mental illnesses. Among these, an internalizing disorder factor (INT) emerged as a robust and reproducible dimension of genetic risk shared among GAD, MDD, and PTSD. This factor was characterized by extensive polygenic overlap and high local genetic correlation, providing strong empirical support for long-standing nosological models of internalizing psychopathology.

While these findings establish the existence of a shared genetic liability underlying internalizing disorders, they leave critical questions unresolved. Factor-analytic models are optimized to capture shared genetic structure but do not directly identify the molecular pathways through which this shared risk operates, nor do they delineate biological mechanisms that may differentiate individual disorders. Due to the limitation of case-case GWAS (CC-GWAS; which is intended for comparing two different disorders with genetic correlation<0.8) approach^2,7^, PGC study could not compare the differences across internalizing pairs. Moreover, enrichment analyses conducted at the factor level necessarily prioritize broad biological processes and may obscure disorder-specific regulatory, cellular, or molecular signatures^8,9^. As a result, the extent to which internalizing disorders are driven by convergent biological mechanisms versus distinct, disorder-specific pathways remains unclear.

Despite the progress made by the PGC cross-disorder study^2^, most genetic studies of internalizing disorders have examined each condition in isolation, limiting our understanding of how much of the genetic risk is shared across GAD, MDD, and PTSD and how much reflects disorder-specific biological pathways. Disentangling these components is essential for clarifying the etiology of internalizing disorders, improving nosological frameworks, and informing the development of targeted prevention and treatment strategies.

Here, we generated novel genome-wide data across GAD, MDD, and PTSD to characterize both shared and disorder-specific genetic architectures. By applying multivariate modeling across different approaches, data types, and perspectives, we identified convergent biological processes underlying internalizing risk and uncovered pathways unique to each disorder.

## RESULTS

We conducted a multivariate and multi-ancestry genome-wide association study of GAD (1,358,762 participants, including 123,006 cases), MDD (3,601,629 participants, including 685,929 cases), and PTSD (1,617,876 participants, including 179,729 cases). These samples represent four population groups: African descent (AFR; N=89,567 for GAD, 260,042 for MDD, 113,651 for PTSD), Admixed-American descent (AMR; N=53,618 for GAD, 436,817 for MDD, 65,871 for PTSD), East Asian descent (EAS; N=7,148 for GAD, 391,113 for MDD, 8,228 for PTSD), and European descent (EUR: N=1,208,429 for GAD, 2,513,657 for MDD, 1,430,126 for PTSD) (Table S1). Comparable heritability estimates and high genetic correlations were observed among the individual cohorts investigated (Table S2). In line with previous evidence^2^, the INT and the three disorder-specific GWASs exhibited a high genetic correlation (rg>0.9) in the European ancestry (Table S3).

### Gene Discovery

We identified 248 genome-wide significant associations (p<5 X 10^−8^) for INT (Figure 1), of which 96 were novel; 109 associations for GAD, with 63 novel loci; 591 associations for MDD, including 241 novel loci; and 237 associations for PTSD, of which 113 were novel (Figure S1-S2, and Table S4). The number of INT lead SNPs in linkage disequilibrium (LD) with disorder-specific lead SNPs was 49 for GAD, 65 for MDD, and 94 for PTSD (Figure 2; Table S4).

**Figure 1:**
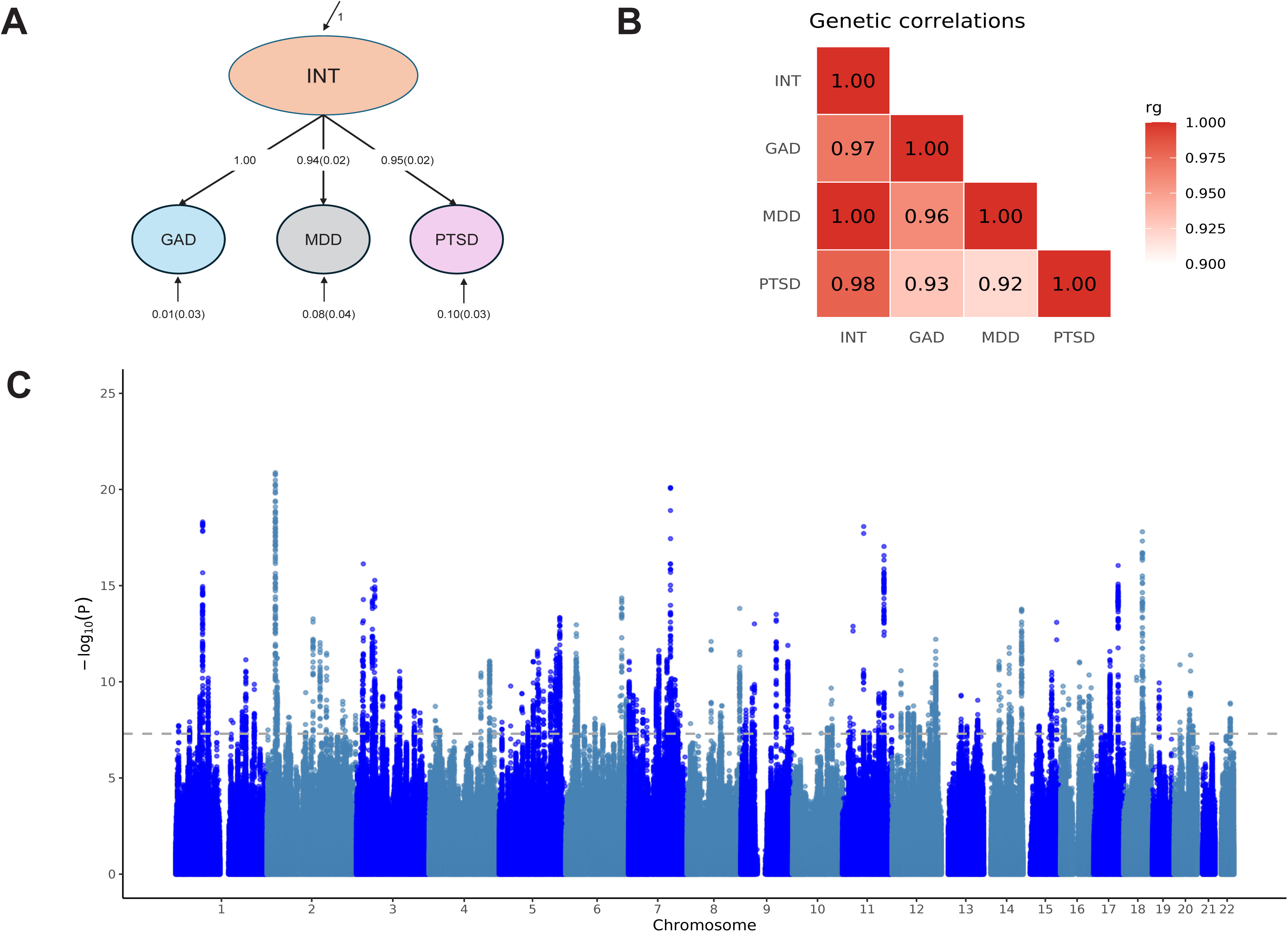
Genomic structural modeling of internalizing disorders. A: Genomic structural model (gSEM) with standardized estimates for internalizing disorder factor (INT); B: Heatmap of genetic correlations (rgs) across INT and disorder-specific traits as estimated using LDSC; C: Manhattan plot of genome-wide association studies for INT by gSEM, the horizontal dashed line represents the genome-wide significance threshold (p=5×10^−8^). INT: internalizing disorder factor; GAD: Generalized anxiety disorder; MDD: Major depressive disorder; PTSD: posttraumatic stress disorder.

**Figure 2:**
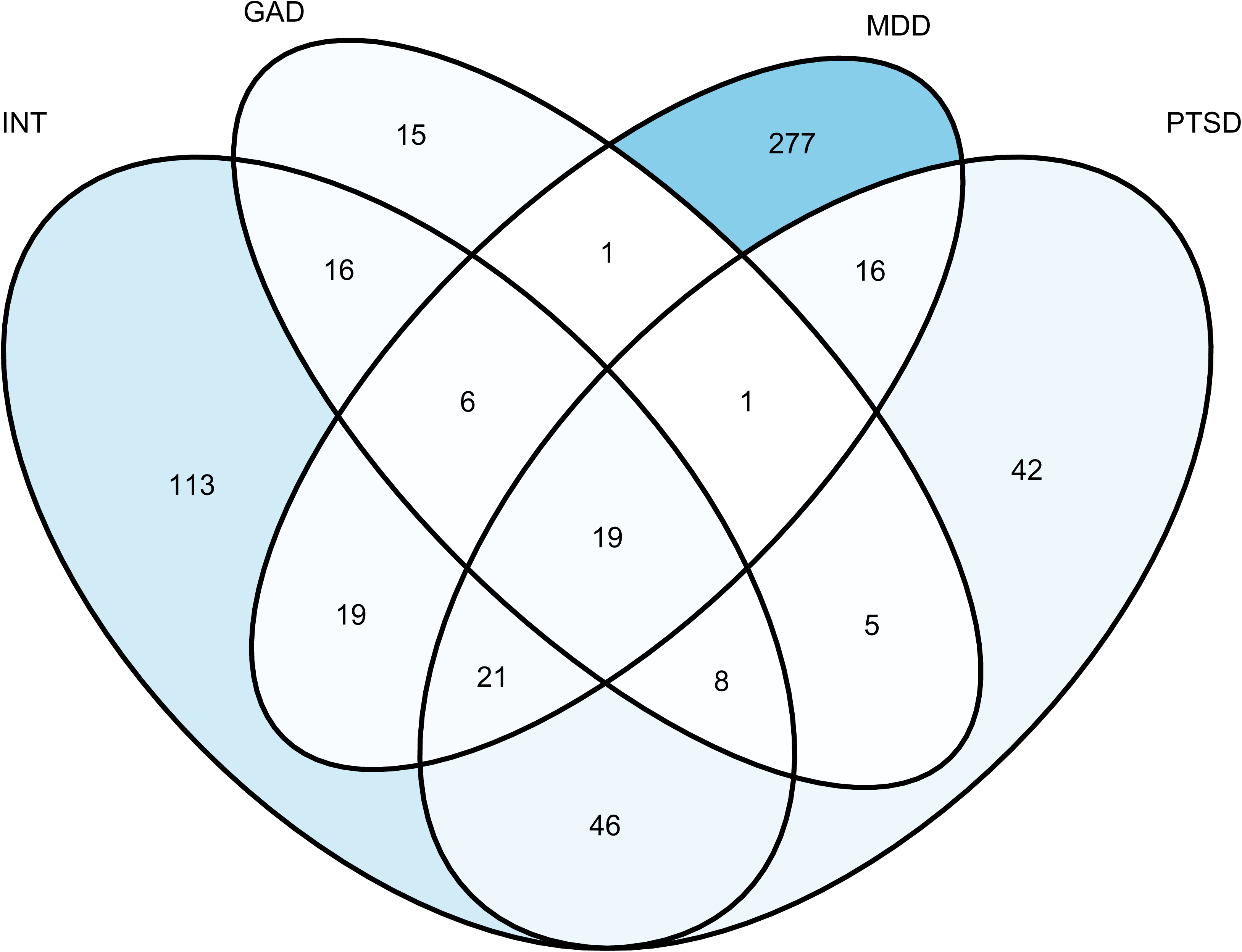
Overlap of shared internalizing disorder factor lead SNPs with disorder-specific lead SNPs in European ancestry. INT: internalizing disorder factor; GAD: Generalized anxiety disorder; MDD: Major depressive disorder; PTSD: posttraumatic stress disorder.

Comparing effect sizes across traits and ancestry groups, we observed several differences, suggesting the presence of ancestry and trait-specific genetic effects (Supplemental Results and Supplemental Table 4). Among the 96 novel INT associations, the most significant variant was rs3020318 (p=4.39×10^−15^), located near *ESR1*. For GAD, we identified 63 novel genome-wide significant loci, of which 14 were unique to EUR ancestry when compared with other traits across ancestries, and 23 were unique to the cross-ancestry GAD GWAS (Table S4B). The most significant GAD variant among the novel associations was rs34092621 (p=3.34×10^−13^ in the cross-ancestry analysis), located near *TMEM110*.

Among the 241 novel MDD loci, 53 were unique to EUR ancestry when compared with other traits across ancestries, and 75 were unique to the cross-ancestry MDD GWAS. The newly identified variant rs1532204 near *BSN* reached genome-wide significance in EUR ancestry (β=0.03, p=9.21×10^−17^) and was also genome-wide significant in the cross-ancestry analysis (Table S4C). For PTSD, we identified 113 novel genome-wide significant loci, including 45 unique to EUR ancestry when compared with other traits across ancestries and 32 unique to the cross-ancestry PTSD GWAS (Table S4D). The most significant PTSD association was rs12025473 (p=7.75×10^−15^ in EUR ancestry), which was also genome-wide significant in the cross-ancestry analysis. With respect to the other population groups, we also identified two novel genome-wide significant associations for AFR GAD (rs144026835, p=2.65×10^−8^; rs370143144, p=2.82×10^−8^), one novel for AMR MDD (rs4665145, p=1.61×10^−8^), two novel for EAS MDD (rs5878866, p=3.23×10^−8^; rs10742983, p=2.82×10^−8^), and one novel for EAS PTSD (rs149517421, p=5.59×10^−9^). No genome-wide significant SNPs were identified for the remaining population groups.

Using GWAS by subtraction approach^10^, we modeled EUR genome-wide association statistics related to GAD, MDD, and PTSD to quantify shared and disorder-specific liability. All three disorders loaded onto the INT factor representing their common genetic variance, while PTSD and MDD additionally loaded onto orthogonal disorder-specific factors (PTSD_SUB and MDD_SUB), capturing residual genetic variance unique to each disorder. Multivariate GWAS of these latent factors identified 67 INT loci, 137 MDD_SUB loci, and one PTSD_SUB locus (Figure S3; Table S5). Overall, GWAS-by-subtraction indicates that all GAD and most PTSD genetic risk is captured by the shared INT factor, whereas roughly 38% of MDD-associated variants reflect disorder-specific mechanisms. Among the unique significant associations (29 of 137 were unique associations, but in LD with lead SNPs from the main MDD GWASs). For INT GWAS by subtraction, 73% of the original signal was attributable to genetic effects shared with these three disorders, while the remaining 27% (67/248) reflects unique genetic contributions to the internalizing disorder factor beyond individual disorders.

Considering LD-independent significant variants in the primary GWAS (r²<0.1), the conditional analysis identified four additional loci for cross-ancestry MDD GWAS, including rs2146372 (pJ=2.34×10^−8^), rs9768784 (pJ=1.78×10^−12^), rs629701 (pJ = 2.70 × 10^−8^), rs145350287 (pJ=3.53×10^−9^). Three additional loci were identified for the EUR MDD, including rs12631293 (pJ=4.53×10^−8^), rs11563286 (pJ = 7.98 × 10^−9^), and rs1474051 (pJ = 4.65 × 10^−8^) (Table S6).

To prioritize plausible causal variants within the genome-wide significant loci identified, we conducted a cross-ancestry fine-mapping analysis using SuSiEx^11^. We identified putative causal evidence (posterior inclusion probability, PIP>0.8) for 455 variants (Table S7). Specifically, SuSiEx identified 53 INT causal SNPs (Table S7A), with three of them showing PIP=1 (i.e., *SOX5* rs12305561, *LINGO1* rs8038666, and *EXOC4* rs12707093). For GAD, we uncovered 15 EUR-causal SNPs and 13 cross-ancestry causal loci (Table S7B). With respect to other population groups, six GAD causal variants were observed in AFR (*TRPV6* rs575403075 and *CER1* rs144026835 not overlapping with EUR GAD loci) and one for EAS (*NEK4* rs34291015 overlapping with EUR-GAD loci).

For MDD, we identified 196 and 185 causal variants in EUR-specific and cross-ancestry fine-mapping analyses, respectively (Table S7C). Eighty-two MDD causal variants overlapped between these analyses. Two variants showed putative causal effects across all four population groups investigated (AFR, AMR, EAS, and EUR): *RNA5SP52* rs1576672 and *WDPCP* rs72813418. Conversely, we also identified two population-specific MDD causal loci: *PLA2R1* rs4665145 in AMR and *GVINP1* rs79708590 in EAS.

For PTSD, 26 putative causal SNPs for EUR and 45 for cross ancestry (Table S7D). Additional population-specific causal loci were observed for AFR (*C10orf11* rs114935459) and EAS (*FOXP1* rs78507955). For MDD_SUB, we identified 62 causal SNPs (Table S7E). Among the genes not overlapping with MDD fine-mapping results (N=19), the top MDD_SUB causal variants (PIP=1) included *IP6K2* rs7610519, *NRM* rs9262146, *HCG20* rs3095335, and *TUBBP9* rs2475811. No PTSD_SUB causal SNP was identified.

### Multi-Omics Characterization

Colocalization analysis of internalizing phenotypes with brain-specific DNA methylation regulation identified 743 SNP-CpG pairs (posterior probability, PP_FC_>0.7, PP_SNP_>0.5), of which 225 (30%) overlapped across different internalizing disorders and factors (Table S8B) in EUR. For instance, *TRIM27* rs3135293 effect on INT and PTSD colocalized with cg06608359 methylation quantitative trait locus (mQTL; PP_FC_=1, PP_SNP_=1), and *FOXP2* rs10228494 association with INT, GAD, and PTSD colocalized with the genetic regulation of cg03504834 (PP_FC_=0.95, PP_SNP_=0.87). We also observed trait-specific mQTL colocalization signals, including *RAB27B* rs2311117 for INT (PP_FC_ = 0.99, PP_SNP_=1), *KLHDC8B* rs4955420 for GAD (PP_FC_=0.99, PP_SNP_=1), and *IK* rs778596 for PTSD (PP_FC_=0.98, PP_SNP_=1). MDD-associated variants showed disorder-specific mQTL colocalization: *TRIM31* rs2023473 (PP_FC_=0.99, PP_SNP_=0.98), *IGSF9B* rs612823 (PP_FC_=0.99, PP_SNP_=0.97), and *EIF4E3* rs36090610 (PP_FC_=0.99, PP_SNP_=0.98).

With respect to the colocalization between genetic associations and brain-specific transcriptomic regulation, we observed multiple instances of colocalization linking pleiotropic effects on INT, GAD, and PTSD to expression QTLs (eQTL; e.g., *CTTNBP2* rs7801876, PP_FC_=0.81, PP_SNP_=0.98; *RGS7BP* rs7719544, PP_FC_=0.78, PP_SNP_=1; *LRFN5* rs2415688, PP_FC_=0.94, PP_SNP_=0.99; Table S8A). Conversely, disorder-specific brain-eQTL colocalization signals were observed with respect to MDD genetic associations (i.e., *SOX21-AS1* rs116988021, PP_FC_=0.79, PP_SNP_=0.53; *LRRK2* rs2638271, PP_FC_=0.77, PP_SNP_=0.90).

Considering colocalization signals shared across brain mQTLs, protein QTLs (pQTL), and single-cell QTLs (scQTL), we observed convergent evidence related to loci associated with multiple internalizing phenotypes as well as trait-specific genetic associations (Table S8). For example, *LRFN5* association with INT, GAD, and PTSD, and *BTN3A2* association with INT, GAD, and MDD, colocalized with brain mQTL, pQTL, and scQTL. Conversely, other loci showed trait-specific colocalization with two or more brain QTL datasets, inlcluding *SLC25A12*, *MDGA2*, and *MUSTN1* for INT; *BTN3A3* and *CCDC92* for GAD; *HLA-C*, *MICB*, and *HCP5* for PTSD; *TNFRSF13C* for MDD.

We identified 193 genes associated with at least one of the internalizing phenotypes after multiple testing correction in both gene-based analyses using multi-marker analysis of genomic annotation (MAGMA)^12^ and pathway scoring algorithm (PASCAL)^13^ (Tables S9 and S10) in EUR. Specifically, MAGMA and PASCAL gene-based results converged 111 genes for INT, 86 for PTSD, 69 for MDD, 39 for GAD, and 11 for MDD_SUB. Among these, the most significant shared genes were *DCC* (MAGMA-p=5.85×10^−18^, PASCAL-p= 1.00×10^−12^) and *ESR1* (MAGMA-p =5.34×10^−13^, PASCAL-p=3.94×10^−7^) for GAD, *FOXP2* for both MDD (MAGMA-p=5.85×10^−18^, PASCAL-p=1.00×10^−12^) and PTSD (MAGMA-p=5.85×10^−18^, PASCAL-p=1.00×10^−12^). *DRD2* was the most significant gene for MDD_SUB (MAGMA-p =3.22×10^−11^, PASCAL-p = 4.66×10^−12^). No gene survived Benjamini-Hochberg correction for PTSD_SUB.

We conducted a multi-tissue transcriptome-wide association study (TWAS) considering 13 brain regions available from GTEx^14^ (Figure 3; Table S11) in EUR. After Bonferroni correction accounting for the 13,457 genes tested (p<3.72×10^−6^), we identified 90 associations with INT (most significant: *RBM6*, p=6.97×10^−16^), 31 with GAD (most significant: *SPCS1*, p=7.52×10^−13^), 67 with PTSD, 57 with MDD, 26 with MDD_SUB, and one with PTSD_SUB (*RP11-344P13.6*, p=2.24×10^−11^). *GPX1* was the most significant genetically regulated transcriptomic association with MDD (p=9.51×10^−18^), PTSD (p=1.35×10^−16^), and MDD_SUB (p=3.19×10^−12^).

**Figure 3:**
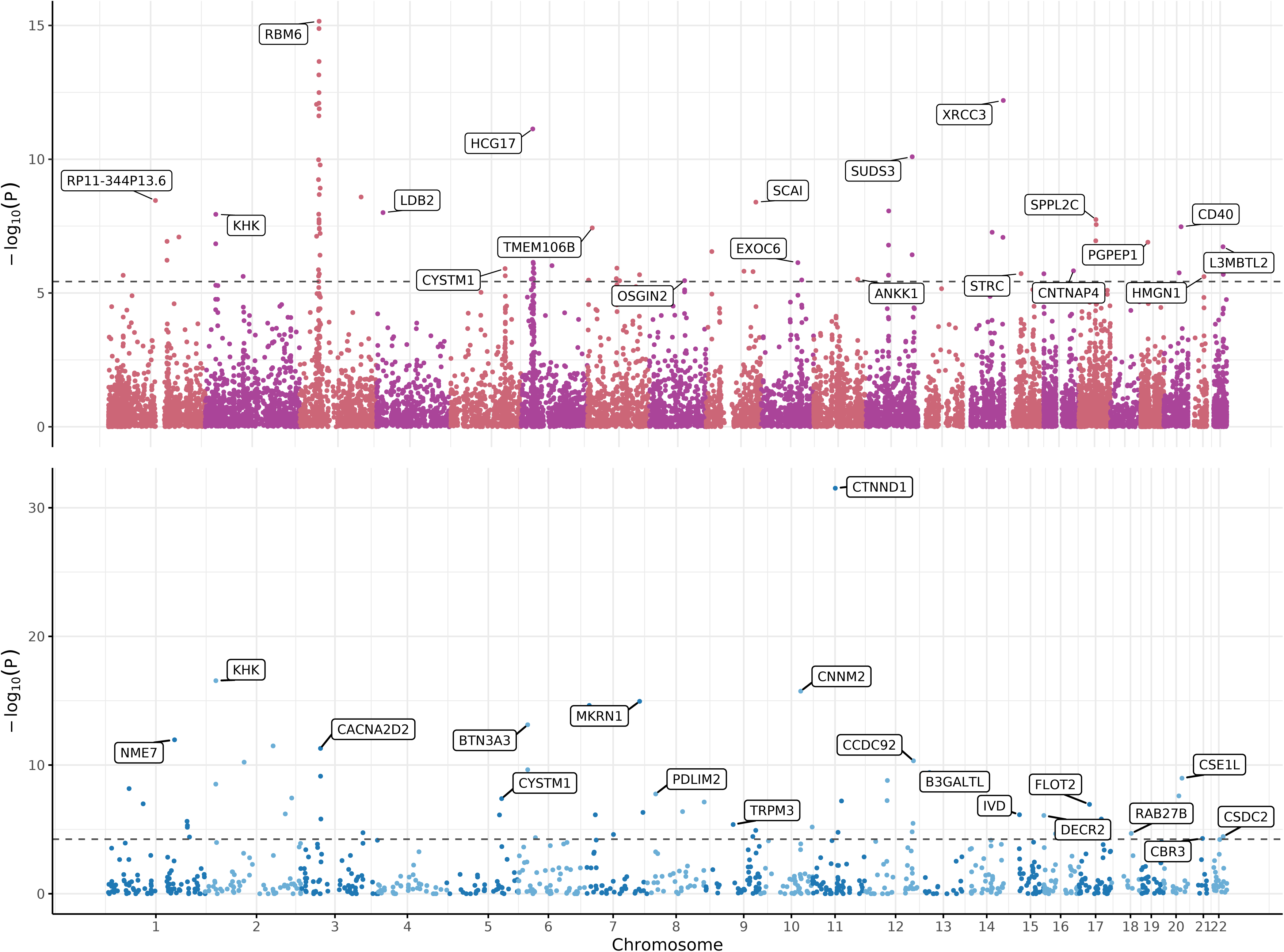
Significant genes identified in the cross-tissue transcriptome-wide association study (TWAS) and proteome-wide association study (PWAS) in dorsolateral prefrontal cortex (DLPC) for the internalizing disorder factor. The upper panel shows significant genes identified in the meta-analysis of cross-tissue TWAS. The lower panel shows significant genes identified in the DLPC-specific PWAS combining the Religious Orders Study and Memory and Aging Project (ROSMAP) and Banner Sun Health Research Institute (Banner) datasets. For visualization clarity, only the most statistically significant gene per chromosome is labeled. Full results are presented in Figures S4-S8.

Our proteome-wide association study (PWAS) for EUR identified 71 Bonferroni-significant protein-coding genes across the internalizing phenotypes (Figure 3, Figure S8; Table S12) in EUR. Specifically, 57 significant associations were detected for INT (most significant: *CTNND1*, p=3.04×10^−32^), while no disorder-specific association survived multiple testing correction. Conversely, we identified 10 genetically regulated proteomic associations with MDD_SUB (most significant: *GMPPB*, p=3.64×10^−14^) and four with PTSD_SUB (most significant: *MTRF1L*, p=6.39×10^−6^).

Summary-based Mendelian randomization (SMR)^15^ integrating brain and blood mQTLs identified a total of 1,432 CpG sites genetically associated with at least one internalizing phenotype investigated after FDR multiple testing correction in EUR, with no evidence of heterogeneity across the mQTL datasets meta-analyzed (Figure S9-S12; Table S13). Among these, five CpG sites were associated with four internalizing phenotypes: *MAD1L1* cg07167778 (GAD p=1.56 X 10^−5^; MDD p=3.28 X 10^−8^; PTSD p=2.24 X 10^−6^; MDD_SUB p=2.76 X 10^−7^), *METTL15* cg08751840 (INT p=4.68 X 10^−8^; GAD p=2.72 X 10^−5^; MDD p=1.12 X 10^−6^; PTSD p=1.85 X 10^−5^), cg12370583 (INT p=3.25 X 10^−6^; GAD p=9.27 X 10^−6^; MDD p=6.82 X 10^−6^; PTSD p=2.02 X 10^−7^), cg12693101 (INT p=1.11X10^−7^; MDD p=1.80X10^−8^; PTSD p=1.62X10^−6^; MDD_SUB p=1.92X10^−6^), and cg20712742 (INT p=5.98X10^−6^; GAD p=2.46X10^−6^; MDD p=6.13X10^−6^; PTSD p=1.51X10^−9^). However, 80% of the CpG sites showed FDR-significant associations with only one of the internalizing phenotypes tested. Among the trait-specific mQTL-SMR results, the most significant CpG association was *RNF123* cg05666287 for INT (p=9.46X10^−40^), *COA8* cg13511324 for GAD (p=4.42X10^−14^), *NCAM1* cg25132257 for MDD (p=2.08X10^−26^), *MIR130A* cg16520038 for PTSD (p=1.88X10^−17^), *ASAH1* cg04912297 for MDD_SUB (p=2.10X10^−9^), and *CCDC63* cg16202755 for PTSD_SUB (p=6.20X10^−11^).

Integrating the complementary approaches used to characterize the loci associated with internalizing phenotypes using multi-omics information, we identified a total of 1,291 genes (Figure 4; Table S14) in EUR. Of these, 40% showed evidence related to two or more internalizing phenotypes. The highest convergence across methods (N=10) and disorders (N=6) were observed for *LRFN5*, *KPNA2*, *DRD2*, and *RNF123* (Table S14), which were associated with all internalizing phenotypes except for PTSD_SUB. We also identified disorder-specific genes supported by multiple methods. For instance, *EIF4E3* and *TRMT61A* relationship with MDD was supported by four methods, with the former also being identified by two methods in the MDD_SUB analysis. *RHOT1* association with PTSD was also supported by four methods. Finally, GAD appeared to be uniquely related to *NISCH*, *SFMBT1*, and *TLR9* in two analyses. *NRXN1* was identified by three MDD_SUB analyses, two MDD analyses, and one INT analysis, but there was evidence linking this gene to GAD and PTSD. Similarly, *HCP5* was observed in two PTSD_SUB analyses, two PTSD analyses, and one INT analysis, but not in any of the analyses performed with respect to GAD and MDD.

**Figure 4:**
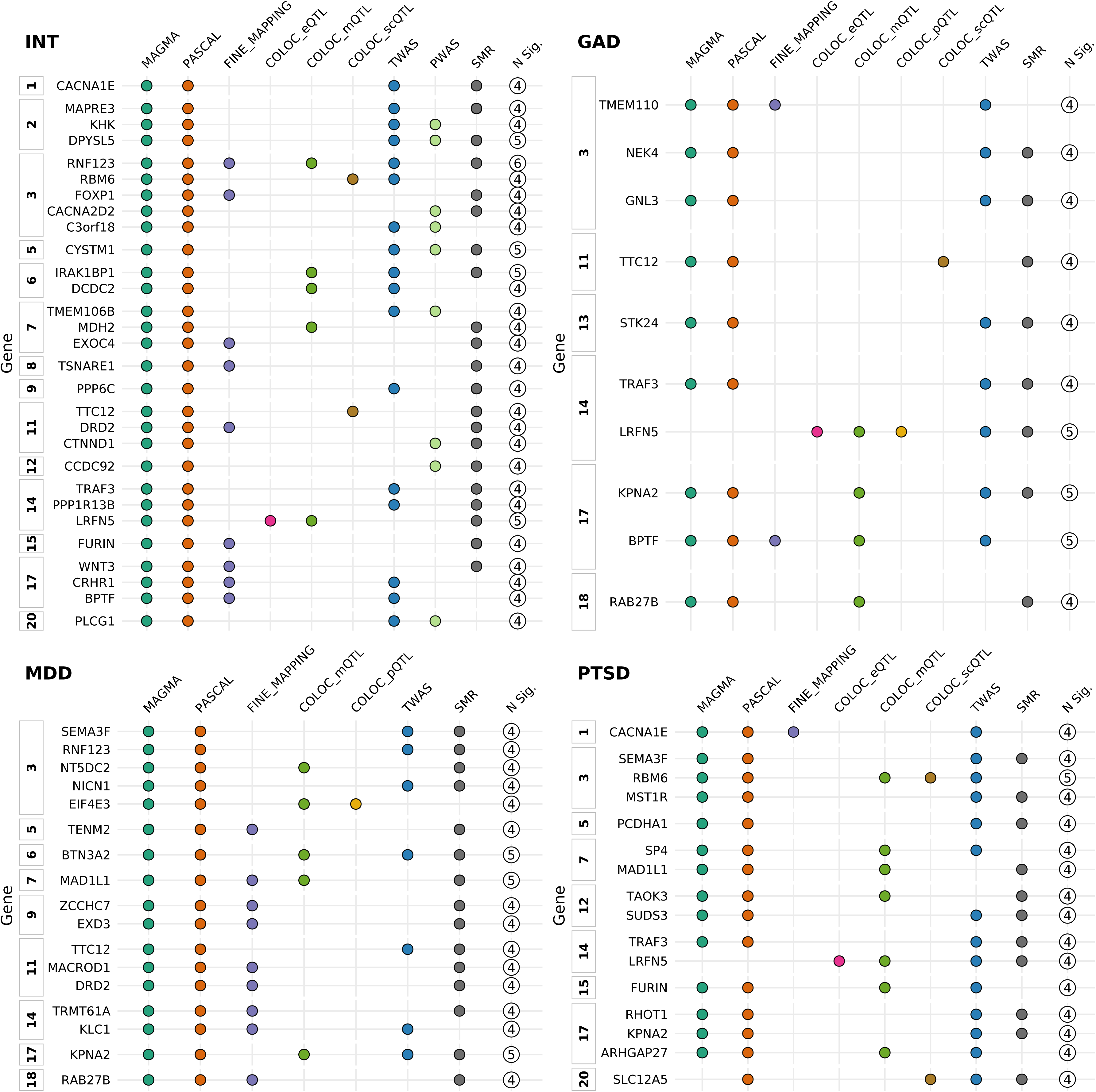
Prioritized Genes integrating evidence from ten gene-discovery analyses across internalizing phenotypes. Upper left panel: Genes with a prioritization score≥4 for internalizing disorder factor (INT) in European descent; Upper right panel: Genes with a prioritization score≥4 for generalized anxiety disorder (GAD) in European descent; Lower left panel: Genes with a prioritization score≥4 for major depressive disorder (MDD) in European descent; Lower right panel: Genes with a prioritization score≥4 for posttraumatic stress disorder in European descent. For each approach and for each internalizing trait, a gene was assigned one point if it showed a significant association in any tissue, dataset, or multi-analysis, and the points were then summed across the six European-ancestry based traits and ten approaches. GAD: Generalized anxiety disorder; COLOC_eQTL: Colocalization analysis with brain cis-expression quantitative trait loci; COLOC_mQTL: Colocalization analysis with methylation quantitative trait loci; COLOC_scQTL: Colocalization analysis with single-cell quantitative trait loci; COLOC_pQTL: Colocalization analysis with brain protein quantitative trait loci; FINE-MAPPING: fine-mapping analysis; INT: internalizing disorder factor; MAGMA: Multi-marker Analysis of GenoMic Annotation; MDD: Major depressive disorder; PASCAL: Pathway Scoring Algorithm; PTSD: posttraumatic stress disorder; PWAS: Proteome-Wide Association Study; SMR: Summary-based Mendelian randomization; TWAS: Transcriptome-Wide Association Study.

### Gene-Ontology Enrichments

In GSA-MiXeR analysis for EUR^16^, 814 gene ontologies (GO) showed SNP-based heritability enrichment for at least one internalizing phenotype after Bonferroni correction accounting for the number of terms tested (p<4.77×10^−6^; Table S15). Among GO enrichments related to INT factor, the most statistically significant were biological processes related to neurogenesis (GO:0022008 p=1.94X10^−167^) and neuron differentiation (GO:0030182 p=2.15X10^−143^). These GO terms were also Bonferroni significant with respect to MDD (p=1.19X10^−121^ and 1.02X10^−70^, respectively), MDD_SUB (p=1.61X10^−70^ and 7.35X10^−56^, respectively), and PTSD_SUB (p=6.70X10^−54^ and 5.75X10^−49^, respectively). Among the top GAD findings (p<10^−300^), we observed the node of Ranvier (GO:0033268), which did not reach Bonferroni-corrected significance with respect to any of the other phenotypes. Cellular component related to synapse (GO:0045202) was among the most significant GO enrichments for INT (p=6.79X10^−130^), MDD (p=1.12X10^−113^), MDD_SUB (p=1.48X10^−82^), and PTSD_SUB (p=3.48X10^−27^). This GO term was also Bonferroni significant with respect to GAD (p=3.23X10^−8^). Interestingly, among phenotype-specific results, microtubule bundle formation (GO:0001578) reached Bonferroni significance only with respect to MDD_SUB (p=3.35X10^−7^) and PTSD_SUB (4.41X10^−6^).

### Drug Repurposing

Two complementary approaches were used to translate genetic findings into candidates for drug repurposing in EUR. The gene2drug approach^17^ leveraged GO enrichments to identify molecular compounds targeting the pathogenesis of internalizing phenotypes, while DRUGSETS^18^ analysis assessed the overlap between gene-based associations and potential therapeutics. GO-based drug repurposing uncovered nine compounds (Supplemental Table 16). Among these, abamectin showed Bonferroni significant enrichment (p<3.82X10^−5^) with respect to INT and PTSD_SUB and suggestive enrichment (p<0.01) with respect to GAD, MDD, PTSD, and MDD_SUB. Fluspirilene was the most significant INT-related molecular compound (p=4.70X10^−7^), with also suggestive evidence (p<0.01) with respect to GAD, PTSD, MDD_SUB, and PTSD_SUB. Lovastatin was the most significant GAD-related drug (p=1.81X10^−7^), although no suggestive convergence was observed with respect to the other internalizing phenotypes investigated (p>0.01). Regarding PTSD, we identified perhexiline (ES=0.32, p=1.76X10^−5^); however, suggestive evidence observed for this drug in the INT analysis showed the opposite enrichment direction (ES=-0.43, p=0.002).

After Bonferroni multiple testing correction accounting for the number of candidates tested (p<6.8×10^−5^), DRUGSETS approach^18^ identified 18 therapeutics converging on the genes associated with internalizing phenotypes (Supplemental Table 17). Among these, etonogestrel was linked to INT (p=1.15X10^−8^), GAD (p=7.99X10^−9^), and MDD (p=1.89X10^−5^). Other compounds related to hormonal regulation were also identified: dehydroepiandrosterone-sulfate (MDD and MDD_SUB), chlorotrianisene (MDD_SUB), clomifene (INT and GAD), desogestrel (INT and GAD), estriol (MDD and MDD_SUB), gestrinone (INT and GAD), hydroxyprogesterone (INT and GAD), levonorgestrel (INT and GAD), and tibolone (MDD and MDD_SUB). We observed several antipsychotic medications: acetophenazine (MDD_SUB), cariprazine (MDD_SUB), fluphenazine-decanoate (MDD and MDD_SUB), and tiapride (MDD and MDD_SUB). Our analysis also uncovered the overlap between internalizing-associated genes and pain medications: dihydroergotamine (MDD), gabapentin (GAD), and rotundine (MDD).

### Pleiotropy Analysis

We applied MiXeR bivariate modeling^19^ to investigate the pleiotropy of internalizing phenotypes with other psychiatric disorders in EUR. Statistically meaningful models permitted us to analyze INT, GAD, MDD, and PTSD with respect to alcohol use disorder (AUD)^20^, bipolar disorder (BIP)^21^, cannabis use disorder (CUD)^22^, and schizophrenia (SCZ)^23^. Overall, similar pleiotropic patterns were observed between internalizing phenotypes and these other mental illnesses, with relatively small differences among INT, GAD, MDD, and PTSD (Figure 5, Table S18). However, BIP showed a distinctive pattern with respect to the phenotypes derived from the GWAS-by-subtraction analysis. Specifically, while MDD and MDD_SUB showed similar genetic correlations (0.50 and 0.44, respectively) and fraction of shared variants (0.64 vs. 0.74, respectively), BIP-PTSD pleiotropy (rg=0.44, shared-fraction proportion=0.51) was much different from the one observed between BIP and PTSD_SUB (rg=0.11, shared-fraction proportion=0.05).

**Figure 5:**
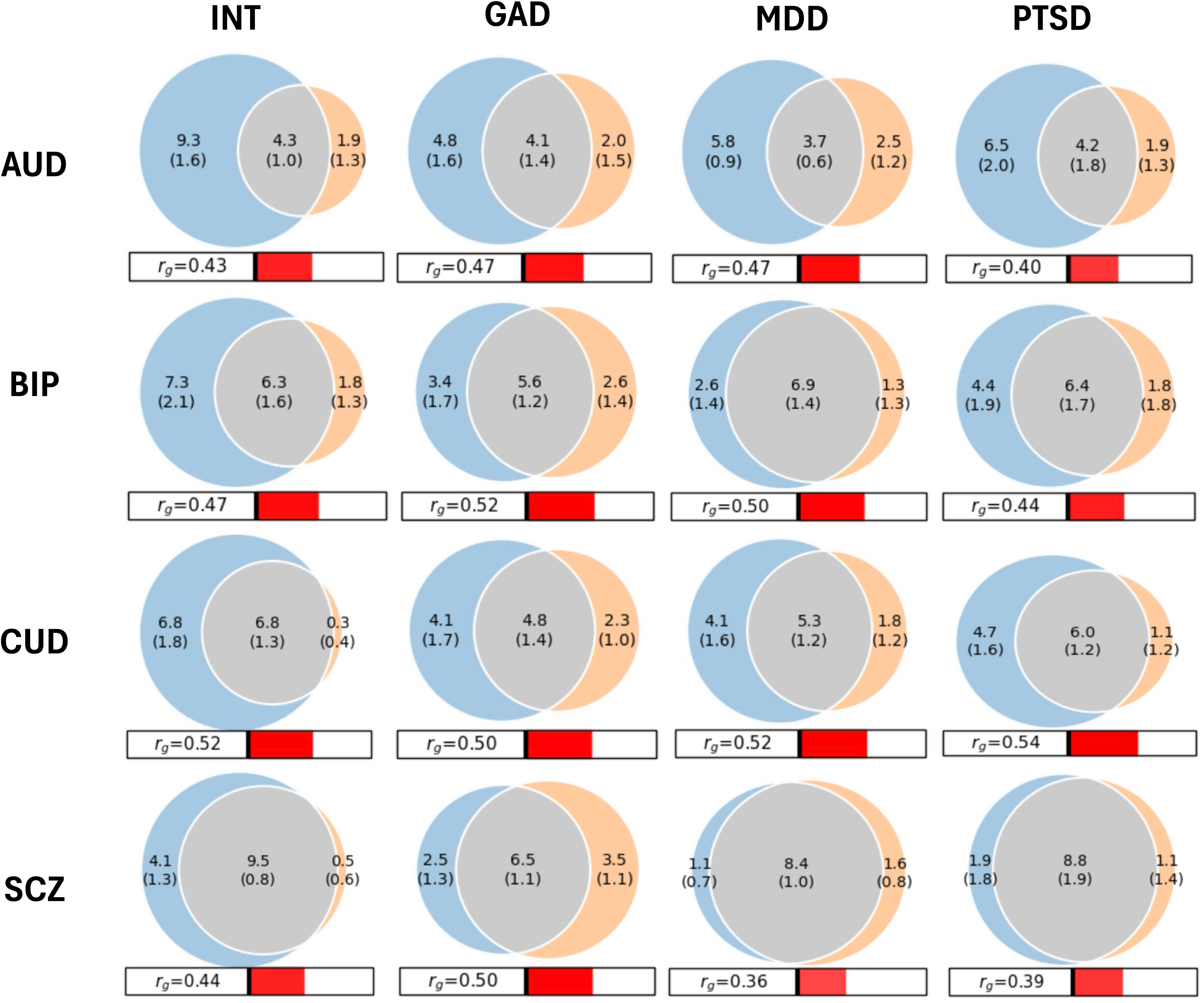
Genetic relationships between internalizing phenotypes and main psychiatric disorders. Venn plots of variants shared between internalizing phenotypes and other main psychiatric disorders. GAD: Generalized anxiety disorder; AUD: Alcohol use disorder; BIP: bipolar disorder; CUD: Cannabis use disorder; INT: internalizing disorder factor; MDD: Major depressive disorder; PTSD: posttraumatic stress disorder; SCZ: schizophrenia.

Through a phenome-wide analysis, we investigated the genetic correlations between the internalizing phenotypes and 11,374 complex traits available from the UK Biobank (UKB)^24^, FinnGen^25^, and the Million Veteran Program (MVP)^26^. Considering GAD, MDD, and PTSD, 1,756 outcomes showed Bonferroni-significant genetic correlations with these three internalizing disorders (p<4.4×10^−6^; Table S19). As expected, top findings included mental health phenotypes such as sertraline prescriptions (GAD rg= 0.92±0.09; MDD rg= 0.99±0.1; PTSD rg=0.88±0.1) and “other reaction to severe stress, and adjustment disorders” (GAD rg=0.96±0.04; MDD rg= 0.94±0.06; PTSD rg=0.89±0.05). However, we also observed high genetic correlations with physical health symptoms and conditions, such as “nausea and vomiting” (GAD rg=0.81±0.1; MDD rg=0.83±0.1; PTSD rg=0.82±0.1), “other ill-defined and unknown causes of morbidity and mortality” (GAD rg=0.82±0.09; MDD rg=0.77±0.08; PTSD rg=0.79±0.09), “constipation” (GAD rg=0.80±0.08; MDD rg=0.81±0.07; PTSD rg=0.75±0.07), “functional digestive disorders” (GAD rg=0.73±0.04; MDD rg=0.75±0.04; PTSD rg=0.71±0.03), and “gastritis and duodenitis” (GAD rg=0.77±0.08; MDD rg=0.68±0.06; PTSD rg=0.71±0.06).

While most of the genetic correlations observed were consistent across internalizing disorders (Table S20), we observed few instances of statistically different rg estimates (difference-p<2.84×10^−5^). Comparing GAD and PTSD, statistically significant differences were present for 20 traits. These were related to internalizing symptoms and definitions, where GAD rg was higher than PTSD rg (e.g., “depression or dysthymia” rg=0.89 vs. 0.85, p=5.34×10^−12^; “any mental disorder” rg=0.91 vs. 0.81, difference-p=1.44×10^−8^; “nervous feelings” rg=0.54 vs. 0.34, difference-p=9.7×10^−8^), and anthropometric traits, where GAD rg was lower than PTSD rg (e.g., body mass index, BMI rg=0.14 vs. 0.22, difference-p=2.57×10^−7^; Leg fat mass rg=0.16 vs. 0.23, difference-p=6.29×10^−6^). Comparing MDD and PTSD, we observe statistically different rg estimates for “Seen doctor for nerves, anxiety, tension or depression” (rg=0.90 vs. 0.78, difference-p=5×10^−8^) and “depression or dysthymia” (rg=0.85 vs. 0.74, difference-p=4.72×10^−6^). While no statistical differences were observed between GAD and MDD after multiple testing correction, the top difference was “nervous feelings” (rg=0.54 vs. 0.40, difference-p=6.53×10^−5^).

To investigate potential causal relationships underlying the genetic correlations observed, we used two complementary approaches: latent causal variable model analysis (LCV)^27^; and generalized summary-data Mendelian randomization (GSMR)^28^. The LCV approach identified 150 phenotypes showing statistically significant genetic causal proportions (GCP, p<7.5×10^−6^) in relation to at least one of the internalizing disorders (Table S20). A GCP>0 indicates a putative causal effect of the internalizing disorder on the phenotype; GCP<0 indicates a putative causal effect of the phenotype on the internalizing disorder. Among them, we observed convergent findings across the internalizing spectrum. Specifically, GAD-MDD shared phenotypes with significant GPC included physical outcomes such as spinal stenosis (GAD GCP=0.76; MDD GCP=0.74), calcific tendinitis (GAD GCP=-0.83; MDD GCP=0.30), and basophil levels (GAD GCP=-0.59; MDD GCP=-0.68).

Similarly, GAD-PTSD shared phenotypes with significant GPC included physical outcomes such as “takes medication for kidney disease without dialysis” (GAD GCP=-0.91; PTSD GCP=-0.74), diseases of the pleura (GAD GCP=-0.74; PTSD GCP=-0.93), and “subarachnoid hemorrhage due to aneurysm” (GAD GCP=-0.20; PTSD GCP=-0.20). MDD-PTSD phenotypes with significant GPC included “age at first episode of depression” (MDD GCP=0.36; PTSD GCP=-0.10) and physical phenotypes such as “cirrhosis of liver without mention of alcohol” (MDD GCP=0.72; PTSD GCP=-0.76) and bundle branch block (MDD GCP=-0.43; PTSD GCP=-0.63).

Considering significant GSMR results (Table S21), we observed convergence with LCV findings in several of the disorder-specific associations. Specifically, both methods showed that the genetic liability to GAD had a putative causal effect on abnormal neurological movements (GCP=0.68, GSMR-beta=0.46), diaphragmatic hernia (GCP=0.79, GSMR-beta=0.40), other laboratory tests (GCP=0.04, GSMR-beta=0.45), peripheral vascular disease (GCP=0.76, GSMR-beta=0.39), and spinal stenosis (GCP=0.76, GSMR-beta=0.57). Similarly, both LCV and GSM analyses highlighted that the genetic liability to MDD potentially affects “acute upper respiratory infections of multiple or unspecified sites” (GCP=0.76, GSMR-beta=0.21) and back pain (GCP=0.46, GSMR-beta=0.16).

## DISCUSSION

In this multivariate and multi-ancestry investigation, we expanded gene discovery, molecular characterization, and pleiotropy assessment of GAD, MDD, and PTSD, highlighting similarities and differences across internalizing disorders. Compared to previous studies^2,4–6^, we greatly increased the number of known loci associated with INT (96 novel loci), GAD (63 novel loci), and PTSD (113 novel loci). With respect to MDD, the number of genome-wide significant associations identified in the present study was lower than the one observed in the most recent PGC GWAS^3^. This is mostly due to the fact that we had access only to a subset of the 23andMe Research Institute data available to PGC investigators and that we applied stricter LD filtering in our conditional analysis. Nevertheless, including diverse participants enrolled in the All of Research Program (AoU)^29^ in our GWAS meta-analysis permitted us to uncover 241 novel loci associated with MDD. Additionally, while the present study still presents an overrepresentation of EUR-descent participants compared to other population groups, the inclusion of the AoU cohort in our GWAS meta-analysis allowed us to increase the representation of these other ancestry groups and identify novel loci in AFR for GAD, in AMR for MDD, and in EAS for MDD and PTSD.

With respect to recent large-scale genome-wide studies investigating internalizing disorders^3–6^ and cross-disorder psychopathology^2^, we also advanced the characterization of disorder-specific genetic risk across the internalizing spectrum. Specifically, our GWAS-by-subtraction revealed that the majority of GAD and PTSD genome-wide risk is shared with the shared INT. In contrast, MDD exhibited a markedly larger disorder-specific component, with approximately 38% of associated variants reflecting genetic effects independent of shared internalizing liability. This may be because the larger sample size available for the MDD GWAS meta-analysis permitted us to identify disorder-specific genetic effects that may have smaller effect sizes than variants shared across internalizing spectrum. In the following sections, we will discuss the novel evidence generated by the present study with respect to similarities and differences in gene discovery, molecular dynamics, and pleiotropy relationships among GAD, MDD, and PTSD.

Among the 455 putative causal SNPs identified through the cross ancestry fine-mapping analysis, we observed a limited number of loci with shared signals across the disorders investigated (PIP>0.8). *TSNARE1* rs13262595 and *MACROD2* rs80044408 were shared between INT and PTSD. Rs13262595 and other *TSNARE1* variants have been associated with psychiatric disorders, including opioid use disorder^30^, SCZ-BIP^31^, sleep disorders^32^, and suicide behaviors^33^. This gene has been previously implicated in the regulation of endosomal trafficking at the dendritic shaft and spines in mature neurons ^34^. Rs80044408 has been previously associated with smoking initiation ^35^, while other *MACROD2* variants have been associated with multiple human traits, including brain functional variation ^36,37^, basal ganglia structure ^38^, educational attainment ^39^, and alcohol drinking ^35^. In brain, *MACROD2* appears to play an important role in neuronal function and synaptogenesis in the hippocampus^40^.

In our fine-mapping analysis, rs71483742 (intergenic) was identified as a shared causal locus for both MDD and PTSD. To our knowledge, no GWAS has previously identified this variant. Because of its intergenic location, further studies will be needed to understand its functional role underlying the association with internalizing disorders. We also observed that *FURIN* rs4702 is a shared causal locus for GAD and MDD. This is in line with multiple studies showing the impact of this variant across psychiatric disorders and traits^8,30,41^. With respect to internalizing disorders, previous evidence highlighted rs4702 impact on *FURIN* regulation of BDNF maturation, which is a known biological mechanism implicated in GAD and MDD and represents a pharmacological target for such disorders^42,43^.

The top fine-mapped loci related to INT (PIP=1) included *SOX5* rs12305561, *EXOC4* rs12707093, and *LINGO1* rs8038666. *SOX5* is involved in early cortical development and neuronal differentiation^44,45^, suggesting a link between neurodevelopmental processes and internalizing disorders. *EXOC4* locus has been previously associated with MDD^46^, cognitive function^47^, and Alzheimer’s disease^48^, highlighting the importance of regulatory processes related to vesicle trafficking, membrane protein delivery, and neurotransmitter receptor localization at synapses^49^.To our knowledge, no previous GWAS identified rs8038666 as a lead variant. However, other *LINGO1* variants have been reported as associated with well-being spectrum^50^ and neuroticism^51^. This suggests that internalizing spectrum may be affected by altered *LINGO1* regulation of neuronal survival, axonal regeneration, and oligodendrocyte differentiation^52^.

Among GAD fine-mapped variants, *HS3ST2* rs189008015 showed the strongest statistical evidence (PIP=1). This variant has been previously associated with SCZ^53^ and other *HS3ST2* variants have been linked to educational attainment^39^, alcohol consumptions^35^, and MDD in trauma-exposed individuals^54^. *HS3ST2* has been implicated in Alzheimer’s disease because of its sulfotransferase enzyme activity^55^. With respect to MDD, seven loci showed PIP=1 in EUR-MDD, cross-ancestry MDD, and MDD_SUB analyses: rs1002655 (intergenic), *ELAVL4* rs11205679, *ROBO2* rs4684031, rs45534736 (intergenic), *GTF2IRD1* rs12534903, *DCC* rs11663393, and *TCF4* rs12967143. Among these, we observed convergent evidence from MAGMA and SMR analyses for *ELAVL4*, a neuron-specific RNA-binding protein critical for synaptic plasticity and stress responsiveness^56,57^. In the PTSD fine-mapping analysis, we identified two causal SNPs with PIP=1: rs13091933 (intergenic) and *PROX1-AS1* rs6540807. The former has been associated with fat body distribution^58^. Rs6540807 is in LD with a novel locus associated with neuroticism^59^. *PROX1-AS1* is a long non-coding RNA that has been observed in transcriptomic analyses of Alzheimer’s disease^60^ and SCZ^61^.

Our integrative multi-omic analyses provided additional evidence for shared genetic mechanisms influencing internalizing-spectrum risk and disorder specific risk through regulation of gene expression, protein abundance, and DNA methylation. In total, 64 genes demonstrated significant associations in both shared and disorder-level analyses (INT, GAD, MDD, PTSD), indicating partially overlapping biological architectures. Among these, 33 genes were supported by significant signals in at least five of the six tested GWAS datasets, highlighting a subset of highly robust cross-phenotype associations. Notably, *LRFN5*, *KPNA2*, *DRD2*, *RNF123*, *MAD1L1*, and *RAB27B* were each supported by signals from numerous analytical frameworks, highlighting them as high-confidence candidates, and span diverse functional categories, including synaptic organization (*LRFN5*)^62^, nuclear transport and cell-cycle regulation (*KPNA2, MAD1L1*) ^63–65^, dopaminergic signaling (*DRD2*)^66^, protein ubiquitination (*RNF123*) ^67,68^, and vesicle trafficking (*RAB27B*) ^69^, consistent with a broad, polygenic architecture underlying internalizing disorders.

In addition to this shared genetic component, we also identified differences in the genetic architecture across phenotypes. The INT factor showed a large set of genetic associations not observed in disorder-specific analyses, indicating that shared liability across the internalizing spectrum is not solely attributable to the aggregation of disorder-level risk loci. In total, 439 genes were significantly associated exclusively with the INT factor. Among these, seven genes (*RANGAP1, STYXL1, POT1, SRPK2, PSMG1, and ATF6B*) were supported by at least three independent multi-omic analyses, providing convergent evidence for their involvement. These genes implicate fundamental cellular processes, including alternative splicing (*SRPK2*)^70^, nucleocytoplasmic transport (*RANGAP1*)^71^, proteostasis (*PSMG1*)^72^, and endoplasmic reticulum stress responses (*ATF6B*)^73^. Although some INT-specific genes lack direct prior associations with psychiatric disorders, their functions intersect with core regulatory mechanisms, such as transcriptional control and genome stability (*STYXL1* and *POT1*)^74,75^ that have been increasingly linked to mental health outcomes. Together, these findings suggest that shared internalizing liability may arise from perturbations in basic cellular homeostasis that broadly influence neuronal function.

Beyond INT-associated loci, our disorder-specific analyses identified functionally specialized genes. For instance, we observed convergent evidence from two or more approaches linking *TRMT61A*, *MARK3*, and *PTPMT1* to MDD. These genes are implicated in translational regulation (*TRMT61A*) ^76^, intracellular kinase signaling and cytoskeletal regulation (*MARK3*)^77^, and mitochondrial phospholipid metabolism (*PTPMT1*)^78^, highlighting the role of cellular processes essential for neuronal energy homeostasis and signal integration. Consistent with these findings, GWAS-by-subtraction further identified 17 genes exclusively associated with MDD_SUB (e.g., *LRRC23*, *PACSIN2*, *ASAH1*), implicating membrane trafficking, cytoskeletal organization, and sphingolipid metabolism, suggesting that MDD-specific genetic risk reflects disruptions in intracellular signaling and metabolic regulation, and broad neurodevelopmental pathways^79–81^.

For GAD, *NISCH*, *SFMBT1*, and *TLR9* were supported by two or more independent analyses. Among these, *SFMBT1* highlights the role of epigenetic regulation in GAD pathogenesis^82^, while *TLR9* supports the importance of immune-brain interactions^83^. With respect to PTSD, the four genes identified by three or more analytical approaches (i.e., *RHOT1*, *CPEB3*, *LRRC37B*, and *PCDHA3*) are implicated in mitochondrial trafficking (*RHOT1*)^84^, activity-dependent mRNA translation (*CPEB3*)^85^, Ras/MAPK signaling (*RASGRP1*)^86^, and neuronal adhesion (*PCDHA3*)^87^. These pathways are central to synaptic plasticity and experience-dependent circuit remodeling, which are key aspects involved in fear learning and extinction^88^. GWAS-by-subtraction further identified 17 genes exclusively associated with PTSD_SUB (e.g., *CDH3*, *FECH*, *RGS1*), implicating cellular adhesion^89^, metabolic processes, and immune signaling^90,91^, indicating that residual, disorder-specific genetic effects persist beyond shared internalizing liability. Collectively, these results support a hierarchical model of genetic risk across internalizing psychopathology. The INT factor seems to capture shared liability driven by broadly acting cellular and regulatory mechanisms, whereas disorder-specific risk reflects more specialized perturbations of neurodevelopmental, synaptic, and stress-responsive pathways.

Our enrichment analysis revealed 815 significant GO terms linked with at least one of the internalizing disorders. As expected, the top-findings converging among multiple internalizing phenotypes were related to brain-related terms such as neurogenesis, neuron differentiation, and synapse. However, we also observed disorder-specific enrichments. For instance, the node of Ranvier (GO:0033268) was among the top findings for GAD, but did not reach Bonferroni significance in any of the other phenotypes investigated. Evidence from animal models suggested that the remodeling of the node of Ranvier contributes to aberrant circuit function associated with anxiety disorders^92^. Interestingly, only MDD_SUB and PTSD_SUB showed Bonferroni-significant enrichment for microtubule bundle formation (GO:0001578). This may support the hypothesis that disorder-specific components may be more related to specialized perturbations.

Our GO-based drug-repurposing analysis identified fluspirilene as a compound targeting molecular pathways shared across internalizing disorders. This antipsychotic medication is a dopamine receptor blocker, which has also demonstrated anxiolytic and mood-elevating properties^93^. Another interesting drug was lovastatin, which was the top result for GAD. In mice, this medication appears to increase neurogenesis and reduce depressive behaviors^94^. Among the potential therapeutics, we also observed fisetin with respect to both GAD (p=1.91X10^−5^) and PTSD (p<0.01). This natural flavonoid appears to mimic antidepressant action by activating TrkB receptors and modulating serotonin/norepinephrine signaling^95^. With respect to other compounds identified in the GO-based drug-repurposing, abamectin showed convergent evidence across multiple internalizing phenotypes. In mice, chronic exposure to abamectin appears to cause spatial memory deficit and depressive behavior in mice^96^.

Applying an orthogonal drug-repurposing approach based on gene-based associations, we identified multiple molecular compounds related to hormonal regulation (e.g., dehydroepiandrosterone-sulfate, chlorotrianisene, clomifene, and desogestrel). This is in line with other studies, which highlight the potential of hormonal medications to prevent and treat anxiety and depressive symptoms^97,98^. This analysis also highlighted several antipsychotic medications (i.e., acetophenazine, cariprazine, fluphenazine-decanoate, and tiapride) in relation to both MDD and MDD_SUB. This suggests further that antipsychotic drugs target pathogenic processes shared between SCZ and MDD^99^. Pain medications (dihydroergotamine, gabapentin, and rotundine) were another drug category we observed, highlighting the potential of targeting pain sensitivity to treat anxiety and depression^100,101^.

In line with the high genetic correlation among GAD, MDD, and PTSD, we observed similar pleiotropy patterns with respect to other psychiatric disorders. However, we observed that PTSD_SUB (i.e., the disorder-specific component of PTSD genetic liability) showed a much lower genetic correlation and proportion of shared causal variant with BIP than PTSD. This difference was not present when considering MDD and MDD_SUB. While there is known comorbidity between BIP and PTSD^102^, our finding suggests that, differently from MDD, this may be more related to the impact of shared external factors (e.g., exposure to traumatic events) and vulnerability to internalizing conditions than PTSD-specific pleiotropic mechanisms. Also, when investigating pleiotropy across other medical domains, we observed consistent GAD, MDD, and PTSD genetic correlations.

In addition to brain-related outcomes, these internalizing disorders showed high genetic overlap with multiple gastrointestinal conditions, e.g., constipation, functional digestive disorders, gastritis, and duodenitis. This is in line with the growing literature highlighting the role of the gut-brain axis in the shared genetic etiology between gastrointestinal outcomes and psychopathology^103^. Beyond similar pleiotropy patterns, we also observed interesting GAD-PSTD differences. With respect to mental health, GAD showed a higher genetic correlation with general psychiatric symptoms than PTSD. This may be related to the fact that GAD symptoms encompass a broader range of mental health signs than PTSD^104^. Interestingly, we observed an opposite trend for obesity-related anthropometric traits (e.g., BMI and leg fat mass), where GAD genetic correlations were statistically lower than the PTSD ones. This supports that PTSD association with obesity, metabolic syndrome, and rapid weight gain^105^ may be due to pleiotropic mechanisms.

To further investigate GAD, MDD, and PTSD pleiotropy, we applied two genetically informed causal inference methods to identify high-confidence relationships. While both approaches identified putative causal effects related to both mental and physical health, we observed convergent disorder-specific findings related to seven phenotypes. Specifically, GAD genetic liability appears to affect abnormal neurological movements, diaphragmatic hernia, laboratory test orders, peripheral vascular disease, and spinal stenosis. A possible causal effect may contribute to the link between GAD and neurological disorders and the increased anxiety symptoms observed among patients affected by functional movement disorders^106^. Similarly, GAD genetic risk may contribute to the increased healthcare utilization^107^ and the worse outcomes of peripheral vascular disease^108^ observed among GAD patients. With respect to spinal stenosis, there is an ongoing debate regarding the potential impact of stress and mental health on disease progression^109^. Conversely, the relationship with diaphragmatic hernia is less clear, as this congenital birth defect is not likely to be affected by GAD genetic liability. With respect to MDD, the potential effect of its genetic liability on respiratory infections is supported by previous genetically informed and observational evidence pointing toward the brain-lung axis^110^. Likewise, previous findings support the relationship of MDD genetic risk and back pain, which may be partially due to shared biological mechanisms related to spinal degenerative disease^101^.

While the present study generated new insights into similarities and differences of the genetic architecture of GAD, MDD, and PTSD, there are three key limitations we need to acknowledge. In our investigation, we combined information across cohorts with different assessment strategies and sociodemographic characteristics. As previously discussed^51,111^, diagnosis information derived from self-reported assessments and electronic health records can be affected by confounders. In particular, misclassification among GAD, MDD, and PTSD may have increased apparent similarities and hidden true differences in the genetic architecture of internalizing disorders. Additionally, while our analyses expanded gene discovery in diverse population groups, the limited number of EUR-descent participants did not permit us to characterize regulatory and pleiotropic mechanisms underlying internalizing disorders that may differ across worldwide populations. Finally, the much larger sample size available for MDD analyses may have altered the comparability with GAD and PTSD. For instance, as mentioned above, the higher MDD-specific genetic component observed could be due to the greater statistical power available for this condition.

In conclusion, our findings highlight the highly polygenic architecture underlying the disorders within the internalizing spectrum. In particular, we observed that while GAD and PTSD genetics strongly overlap with INT factor, a notable proportion of MDD loci reflects disorder-specific mechanisms. Nevertheless, by integrating multiple sources, data types, and methodological approaches, we produced a robust set of findings that uncovered similarities and differences at multiple levels, including loci, regulatory dynamics affecting other omic domains, and pleiotropic relationships linking internalizing disorders with other complex traits. Overall, these results advance our understanding of internalizing psychopathology and offer a foundation for future studies exploring rare variants, single-cell and spatial transcriptomics, and integrative causal networks analyses.

## METHODS

### Study Populations

We conducted a multivariate and multi-ancestry GWAS of internalizing disorders, leveraging multiple cohorts and data sources. In particular, we analyzed genome-wide association statistics available from MVP^26^, PGC^112^, iPSYCH^113^, FinnGen^25^, and 23andMe Research Institute^114^ and individual-level data available from UKB^115^ and the All of Us Research Program (AoU)^29^. These previous resources were approved by the relevant institutional review boards and all participants enrolled provided written informed consent. In total, our study investigated 1,358,762 participants (including 123,006 cases) with respect to GAD, 3,601,629 participants (including 685,929 cases) with respect to MDD, and 1,617,876 participants (including 179,729 cases) with respect to PTSD (Table S1).

AoU is an ongoing project aiming to recruit more than one million participants representative of the diversity of the US population.^29,116^ To date, whole-genome sequencing data are available for 414,840 AoU participants. Considering electronic health records (EHR) standardized using the Observational Medical Outcomes Partnership Common Data Model (OMOP CDM), GAD (Concept ID: 442077), MDD (Concept ID: 4152280), PTSD (Concept ID: 436676) were identified from the domain “Conditions”. We also use data from “Personal and Family Health History” survey domain to identify MDD (Concept Id: 1384656) and PTSD (Concept Id:1384443). To maximize our sample size, we further included data from “Emotional Health History and Well-Being” survey domain, using PHQ-9 and GAD-7 identified cases for MDD and GAD.

UKB is a large population-based prospective cohort comprising >500,000 individuals recruited between 2006 and 2010 across England, Wales, and Scotland.^115,117^ At baseline, participants completed extensive web-based questionnaires and underwent physical, cognitive, and biological assessments. In the present study, we conducted a GWAS with respect to GAD (10,579 cases and 376,046 controls). Details regarding GAD analysis in UKB can be found in our previous work.^5^ No MDD and PTSD analyses were conducted in UKB, because UKB data were already used in the genome-wide association analyses conducted for these traits by PGC.

PGC is an international collaborative effort involving over 800 investigators from more than 150 institutions across 36 countries, dedicated to advancing biologically, clinically, and therapeutically meaningful genetic discovery in psychiatric disorders. We leveraged publicly available, ancestry-specific PGC GWAS statistics and combined results across ancestries when appropriate.^3,4,118,119^. For MDD, the PGC GWAS comprised a total of 470,555 cases and 1,945,609 controls across EUR, AFR, EAS, and AMR ancestries. For PTSD, the PGC GWAS included 150,760 cases and 1,129,668 controls spanning EUR, AFR, and AMR ancestry groups. For GAD, the PGC GWAS included 5,761 cases and 11,765 controls of EUR.

Additional GWAS data were obtained from MVP, a large observational cohort of the United States Department of Veterans Affairs (VA) healthcare system that has enrolled over one million racially and ethnically diverse participants^26^. The GWAS statistics used in the present study were generated from an analysis of 24,448 and 175,163 participants of AFR and EUR descent, respectively.^120^ In MVP, GAD was defined as a quantitative phenotype based on the total score of GAD-2 scale. We further incorporated data from FinnGen, a large-scale Finnish research initiative integrating genome-wide genotype data with nationwide health registry information.^25^ From FinnGen release 12, we used GWAS statistics for anxiety disorders (KRA_PSY_ANXIETY), comprising 56,552 cases and 362,304 controls of EUR descent. Data from the iPSYCH cohort^113^ (12,655 cases and 19,225 controls) were also included in our GAD GWAS meta-analysis. Finally, we incorporated MDD genome-wide association statistics (101,168 cases and 750,116 controls) available from the 23andMe Research Insitute.^114^

### Ancestry-Specific and Cross-Ancestry Meta-Analyses

Before meta-analyzing different datasets, the following quality control steps were applied: minor allele frequency>1%, imputation INFO score>0.6 when available, and exclusion of non-rs IDs and non-biallelic variants. For the cross-ancestry meta-analysis, we used METAL^121^ (version 67) to combine GWAS results across cohorts. Because MDD data consisted of only case-control cohorts, we applied the inverse-variance weighted approach, which uses effect size estimates and their standard errors. Conversely, because GAD and PTSD data included both binary and quantitative information, we used the sample-size based approach, which combines p-values and the direction of effect weighted by sample size to account for possible sample overlap. To model the genetic liability across internalizing phenotypes, we then used genomic structural equation modeling (gSEM).^122^ This permitted us to identify a single factor underlying the genetic liability shared across internalizing disorders. All GWAS summary statistics were processed using the munge function prior to running the multivariable version of LDSC used as input to gSEM. The munge function aligns GWAS effects to the same reference allele and restricts to HapMap3 SNPs and SNPs with INFO > 0.9. As recommended by the gSEM developers, the loading of the first GWAS was fixed to 1 for model identification. Since gSEM smooths the non-positive-definite covariance matrix and calculates Zsmooth for the largest difference between pre- and post-smoothed matrices, we retained only variants with Zsmooth<0.025 and PQ (heterogeneity test) ≥ 0.01.

LD score regression (LDSC) metrics, including SNP-h^2^, lambda, mean chi-squared statistic, LDSC intercept, and confounding ratio, were calculated for each meta-analysis^123^. LD scores were calculated using HapMap 3 variants ^124^ and the 1000 Genomes Project EUR reference population.^125^ The LDSC analyses were limited to EUR due to the limited sample size of the GWAS available for the other population groups. To identify independent genome-wide significant loci (p<5×10^−8^), we performed LD-based clumping using PLINK v1.9, applying a 250Ξkb physical distance window and an LD threshold of r²Ξ<Ξ0.1 based on 1000 Genomes reference data matching the ancestry groups investigated in the present study. Independent lead SNPs were defined as the most significant variants within each LD block. In addition to LD-based filtering (r²<0.1), we also applied a ±1Mb window to determine novel SNPs more stringently by comparing with previously reported independent SNPs associated with internalizing disorders^2–5^. LD overlap between INT lead SNPs and lead SNPs for GAD, MDD, and PTSD was assessed using PLINK v1.9 with the 1000 Genomes Project European reference panel^125^, retaining SNP pairs within 1 Mb and r² ≥ 0.6.

### GWAS-by-subtraction

To estimate the disorder-specific components unique to each of the internalizing disorders investigated, we used Genomic SEM implemented in R (v4.2.0) to jointly analyze the EUR GWAS meta-summary statistics for GAD, MDD, and PTSD^10^. Initially, the model included three latent variables representing the shared factor (INT) and disorder-specific factors for PTSD (PTSD_SUB), MDD (MDD_SUB), and GAD (GAD_SUB). A SNP-free model using LDSC-derived genetic covariance matrices showed that GAD was entirely captured by the shared internalizing disorder factor, with no residual disorder-specific signal. Accordingly, GAD_SUB was excluded from further analysis, and the model was rebuilt with only INT, PTSD_SUB, and MDD_SUB. In the full model, each SNP was simultaneously regressed on these latent variables to estimate both shared and disorder-specific genetic effects. This framework allowed each SNP to influence the INT and the disorder-specific factors (PTSD_SUB and MDD_SUB) independently. Similarly to other LDSC-based analyses, this was performed only in the EUR data, because of the limited GWAS sample size available for other population groups. To identify independent genome-wide significant loci (p<5×10^⁻^), we performed LD-based clumping using PLINK v1.9, applying a 250Ξkb physical distance window and an LD r²Ξ< 0.1 based on EUR reference populations available from the 1000 Genomes Project^125^.

### Multi-SNP-Based Conditional & Joint Association Analysis

We performed a joint analysis using GCTA-COJO^126,127^ to identify secondary association signals within the loci reaching genome-wide significance with respect to cross-ancestry and EUR-specific analyses. A stepwise model selection procedure was applied to identify independently associated SNPs. The default parameters were used: 10 Mb for LD distance; 0.9 for multiple regression R^2^ cutoff for collinearity; 0.2 for difference in allele frequency between GWAS and LD reference. SNPs with joint-analysis p-value (pJ)<5×10^−8^ were considered genome-wide significant. Among the SNPs identified by COJO, we further excluded those with LD r²≥0.1 with any lead SNPs identified in the primary GWAS analyses.

### Functional Characterization

To identify putative causal variants underlying genome-wide significant associations, we performed fine-mapping using SuSiEx^11^. Fine-mapping was conducted for loci associated with INT, GAD, MDD, and PTSD. For each locus, the genomic region was defined as ±250 kb around the lead SNP, and the corresponding LD matrix was extracted from the 1000 Genomes Project phase 3 ancestry-specific reference panel matching the population groups investigated in the present study. For the cross-ancestry analyses, the LD matrix from the 1000 Genomes Project EUR reference panel^125^ was used, because this population group represents the majority of the samples investigated.

Using HyPrColoc,^128^ we conducted colocalization analysis to identify genetic variants causally associated with internalizing phenotypes through transcriptomic, proteomic, and methylation regulatory mechanims. The GWAS colocalization with cis-regulatory effects was investigated within ±500kb surrounding the index variants identified by the GWAS performed. If multiple genome-wide significant associations were located within the same genomic region, we prioritized the ±500kb region surrounding the index variants with the most significant association. However, the index variants identified by GWAS-by-subtraction (i.e., MDD_SUB and PTSD_SUB) were investigated in the colocalization analysis, also when index variants related to other internalizing phenotypes (i.e., INT, GAD, MDD, and PTSD) were present in the same region. Colocalization between GWAS signals and molecular QTLs was investigated across 362 genomic regions. We derived brain eQTLs from Genotype-Tissue Expression GTEx V8^129^, brain mQTLs^130,131^, brain pQTL from the Religious Orders Study and Memory and Aging Project (ROSMAP) and Banner Sun Health Research Institute (BSHRI)^132^, and brain single-cell eQTLs from PsychENCODE2^133^.

MAGMA (version 1.10)^12^ was used to perform gene-based association and enrichment analyses. In the pre-processing step, each SNP was mapped to a specific gene. A multiple regression approach was used to properly incorporate linkage disequilibrium LD information between markers and to detect multi-marker effects. To confirm further gene-based associations, we also used PASCAL software^13^, which computes gene scores by aggregating the SNP and p-values using the sum of chi-squared statistics. By employing a default ±50Ξkb window of corresponding gene transcription start sites, each single SNP was mapped to genes. PASCAL allows up to 3000 SNPs per gene by default. EUR populations available from 1,000 Genomes Project^125^ were used as LD refence panel in both MAGMA and PASCAL analyses.

Tissue-specific and cross-tissue TWAS of internalizing phenotypes were performed using S-PrediXcan and S-MultiXcan ^134,135^. Leveraging GTEx v8 data ^14^, tissue-specific predictive models of gene expression and their corresponding LD covariance matrices were previously calculated applying elastic net regressions to account for possible inflation due to polygenicity of traits investigated^136^. Tissue-specific TWAS were limited to 13 brain regions available from GTEx v8. Bonferroni multiple testing correction was applied to account for the number of genes tested (N=13,457; p<3.72×10^−6^).

We conducted a PWAS to identify loci whose cis-regulated protein abundance in the dorsolateral prefrontal cortex (DLPFC) is associated with internalizing phenotypes. This analysis was performed using FUSION ^137^. Following the developers’ recommendations, we employed elastic-net models due to their superior predictive performance compared with other modeling approaches. The PWAS was performed only using EUR data, because of the limited sample size of other population groups. We used cis-pQTL weights trained on postmortem brain tissue from ROSMAP (n=330)^138^ and the BSHRI (n=149)^139^. We performed a PWAS with respect to each internalizing phenotype and pQTL dataset. EUR populations available from the 1000 Genomes Project^125^ were used as LD reference panel. To maximize the PWAS statistical power, we meta-analyzed the ROSMAP and BSHRI results generated with respect to each phenotype. This meta-analysis was conducted using the fixed-effects inverse-variance weighting approach. Only proteins available in both pQTL datasets were considered (N=870). A Bonferroni multiple testing correction was applied to account for the number of loci tested with respect to each phenotype (p<5.74×10^−5^).

We used SMR approach^15^ to evaluate the potential impact of genetically regulated DNA methylation on internalizing disorders. The analysis integrated GWAS data with blood and brain mQTL datasets. Blood-based mQTL resources were integrated from multiple large-scale studies, including the GoDMC catalog^140^, ARIES project^141^, and published datasets from Hatton et al.^142–144^, and McRae et al^145^. Brain-specific SMR analyses were conducted using mMeta-brain mQTL data^130^, derived from the DLPFC (ROSMAP data)^146^, non-dissected fetal brain^131^, and frontal cortex of adult brain^147^. EUR populations available from the 1000 Genomes Project^125^ were used as LD reference panel. We applied standard SMR parameters, including cis-mQTL selection within ±2 Mb, a genome-wide significance threshold of p < 5 × 10^⁻^Ξ, LD pruning (r²=0.05-0.9), and application of the HEIDI test (p>0.05) with up to 20 leading cis-SNPs. Using METAL, we performed a sample size-weighted meta-analysis with genomic control and Z-statistic overlap adjustment^121^ to combine information derived across mQTL datasets. Results surviving FDR multiple testing correction and without evidence of heterogeneity across mQTL datasets (heterogeneity-p>0.05) were considered statistically significant.

To characterize the pathogenesis of internalizing phenotypes, we performed a gene-set enrichment analysis using the GSA-MiXeR framework^16^. This allows us to quantify h^2^ enrichments with respect to predefined gene sets while accounting for LD structure^16^. The primary GSA-MiXeR analysis was performed one with respect to EUR GWAS, because of the limited sample size of other population groups. EUR populations available from 1,000 Genomes Project^125^ were used as LD reference panel. Gene sets were defined based on GO terms, including biological processes, molecular functions, and cellular components. Bonferroni correction was applied to account for the number of GOs tested (N=10,461; p<4.77×10^⁻6^). To reduce redundancy in GO term enrichments and identify representative functional themes, we employed REVIGO^148^ using a semantic similarity cutoff of 0.7 and UniProt as the reference database.

### Drug Repurposing

To identify potential therapeutic candidates for internalizing phenotypes, we performed drug repositioning using two complementary methods. Gene2drug^17^ leveraged pathway-level perturbation profiles derived from Connectivity Map data to identify drugs that may up- or down-regulate pathways associated with internalizing liability. Bonferroni-significant GO terms from GSA-MiXeR and REVIGO were used as input, ensuring robust and semantically coherent pathway representation. In parallel, DRUGSETS drug-gene sets^149^ were assessed using MAGMA competitive gene-set analysis to evaluate enrichment of genetic signal among drug targets. Drug sets were further grouped by clinical indication (IND), Anatomical Therapeutic mechanism of action (MOA), and Classification (ATC) III code, and multiple linear regression on MAGMA t-statistics estimated the influence of drug group membership.

### Pleiotropy Analysis

We performed pairwise MiXeR^19^ analyses between each of the internalizing phenotypes (INT, GAD, MDD, PTSD, MDD_SUB, PTSD_SUB) and six psychiatric traits (AUD^20^, BIP^21^, CUD^22^, obsessive-compulsive disorder (OCD)^150^, opioid use disorder (OUD)^30^, SCZ^23^) to estimate polygenic overlap. These traits were selected based on the statistical power of the corresponding GWAS.

We first applied univariate MiXeR analysis for each of the traits to estimate the trait polygenicity, defined as the number of SNPs accounting for 90% of SNP-heritability. We then performed bivariate mixture model analysis for each trait pair to estimate the shared polygenic component. This approach estimates the number of jointly causal variants independently of effect-direction concordance and further partitions the genetic correlation into shared-component and overall genome-wide contributions. To focus only on statistically meaningful results, we considered only models with positive minimum or maximum AIC values.

We conducted a phenome-wide genetic correlation analysis to quantify GAD, MDD, and PTSD pleiotropy with a wide range of phenotypes available from MVP ^26^, FinnGen^25^, and UKB ^24^. The analysis was limited to traits with robust SNP-based heritability (SNP-h^2^ Z>4). Genetic correlations were estimated using LDSC^123,151^, which evaluates the extent to which SNP-based heritability is shared between two traits. Bonferroni correction was applied to account for the number of traits (N=11,374) tested across the three cohorts. To avoid bias from sample overlap, participants from the analyzed cohort (e.g., UKB participants were excluded when analyzing UKB phenotypes) were excluded from the INT GWAS summary statistics.

### Causal Inference Analysis

To assess whether the genetic correlations between internalizing phenotypes and other complex traits are due to possible causal relationships, we used two genetically informed approaches. First, LCV analysis^27^ estimates the GCP while accounting for sample overlap between the two traits. In the present study, GCP>0 indicates a putative causal effect of the internalizing disorder on the phenotype, while GCP<0 indicates a putative causal effect of the phenotype on the internalizing disorder. We used pre-computed LD scores based on the 1000 Genomes Project Phase 3 EUR data when evaluating GCP estimates. The statistically significant threshold of GCP was defined using Bonferroni multiple testing correction accounting for the total number of traits investigated (N=6,006).

Then, GSMR2^28^ was applied to further estimate potential causal relationships for pairs identified in LDSC genetic correlation analysis. The analysis was performed in a bidirectional strategy. To avoid sample overlap, we redid the meta-analysis and gSEM analysis for internalizing disorders, excluding GWAS data from the same cohort. Randomly sampled UKB EUR genotype data (N = 5,000) were used as the LD reference panel. We used a significance threshold of p<1.0×10^−6^ to select instrumental variables (IV) for both internalizing disorders and phenome-wide complex traits. For GWAS with more than 1,500 IVs, we restricted it to the top 1,500 variants. Other parameters were set as follows: multi_snps_heidi_thresh = 0.01, nsnps_thresh = 5, ld_r2_thresh = 0.05, ld_fdr_thresh = 0.05, and gsmr2_beta = 1. In all analyses, Bonferroni correction was applied to account for 6,006 tests across Pan-UKB, Finngen, and MVP data. To avoid bias from sample overlap, participants from the analyzed cohort (e.g., UKB participants were excluded when analyzing UKB phenotypes) were excluded from the INT GWAS summary statistics.

## DATA AVAILABILITY

The GWAS summary statistics generated in the present study will be made publicly available online upon publication. Individual-level data were obtained from UKB (http://www.ukbiobank.ac.uk/) and the AoU Curated Data Repository version 8 (https://www.researchallofus.org). GWAS data used in this study were downloaded from Pan-UKB (https://pan.ukbb.broadinstitute.org/), FinnGen (https://www.finngen.fi/en/access_results), MVP (https://dbgap.ncbi.nlm.nih.gov/); PGC (https://www.med.unc.edu/pgc/download-results/); iPSYCH (https://ipsych.dk/en/research/downloads); 23andMe Research Institute (https://research.23andme.com/publications/).

## CODE AVAILABILITY

The study used the following computational packages: METAL, https://github.com/statgen/METAL; PLINK, https://www.cog-genomics.org/plink/; gSEM, https://github.com/GenomicSEM/GenomicSEM; GWAS-by-Subtraction, https://github.com/PerlineDemange/non-cognitive/tree/master/GenomicSEM/Cholesky%20model; GCTA-COJO, https://yanglab.westlake.edu.cn/software/gcta/#COJO; SuSiEx, https://github.com/getian107/SuSiEx; HyPrColoc, https://github.com/cnfoley/hyprcoloc; MAGMA, https://ctg.cncr.nl/software/magma; PASCAL, https://www2.unil.ch/cbg/index.php?title=Pascal; LDSC, https://github.com/bulik/ldsc; MetaXcan, https://github.com/hakyimlab/MetaXcan; FUSION, https://github.com/gusevlab/fusion_twas; SMR, https://yanglab.westlake.edu.cn/software/smr/; MiXeR, https://github.com/precimed/mixer; GSA-MiXeR, https://github.com/precimed/gsa-mixer; REVIGO, http://revigo.irb.hr/; gene2drug, https://gene2drug.tigem.it/index.php; DRUGSETS, https://github.com/nybell/drugsets; LCV, https://github.com/lukejoconnor/LCV; GSMR2, https://github.com/JianYang-Lab/gsmr2.

## ACKNOLEDGMENTS

The authors acknowledge support from the National Institutes of Health (RF1 MH132337 to R.P.), the American Foundation for Suicide Prevention (PDF-0-065-23 to J.H.), and the MQ Foundation (UFA21\100014 to B.C.M.). We also acknowledge the contribution of the participants and the investigators involved in the UKB, the FinnGen Project, the MVP, the AoU Research Program, and all included studies in the PGC GWAS. We would like to thank the research participants and employees of 23andMe Research Institute for making this work possible. The research using UKB resources has been conducted under Application Number 58146. The AoU Research Program is supported by the National Institutes of Health, Office of the Director: Regional Medical Centers: 1 OT2 OD026549; 1 OT2 OD026554; 1 OT2 OD026557; 1 OT2 OD026556; 1 OT2 OD026550; 1 OT2 OD 026552; 1 OT2 OD026553; 1 OT2 OD026548; 1 OT2 OD026551; 1 OT2 OD026555; IAA: AOD 16037; Federally Qualified Health Centers: HHSN 263201600085U; Data and Research Center: 5 U2C OD023196; Biobank: 1 U24 OD023121; The Participant Center: U24 OD023176; Participant Technology Systems Center: 1 U24 OD023163; Communications and Engagement: 3 OT2 OD023205; 3 OT2 OD023206; and Community Partners: 1 OT2 OD025277; 3 OT2 OD025315; 1 OT2 OD025337; 1 OT2 OD025276.

