## Supplementary Figures for "Multivariate genome-wide association study dissects shared biology and disorder-specific loci across internalizing spectrum in millions of ancestrally diverse participants"

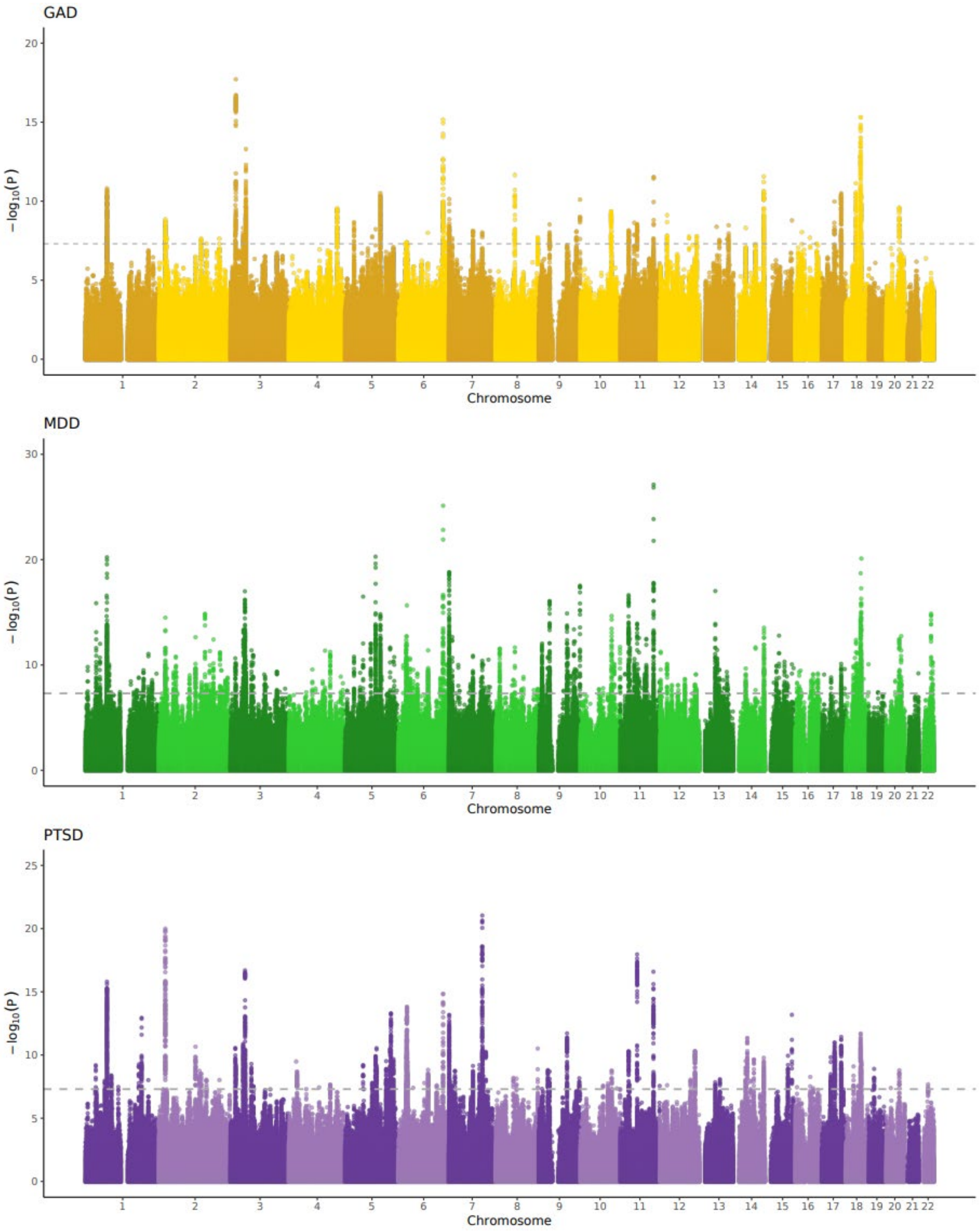
GAD
MDD
PTSD
−log10(P)
Chromosome
1 2 3 4 5 6 7 8 9 10 11 12 13 14 15 16 17 18 19 20 21 22

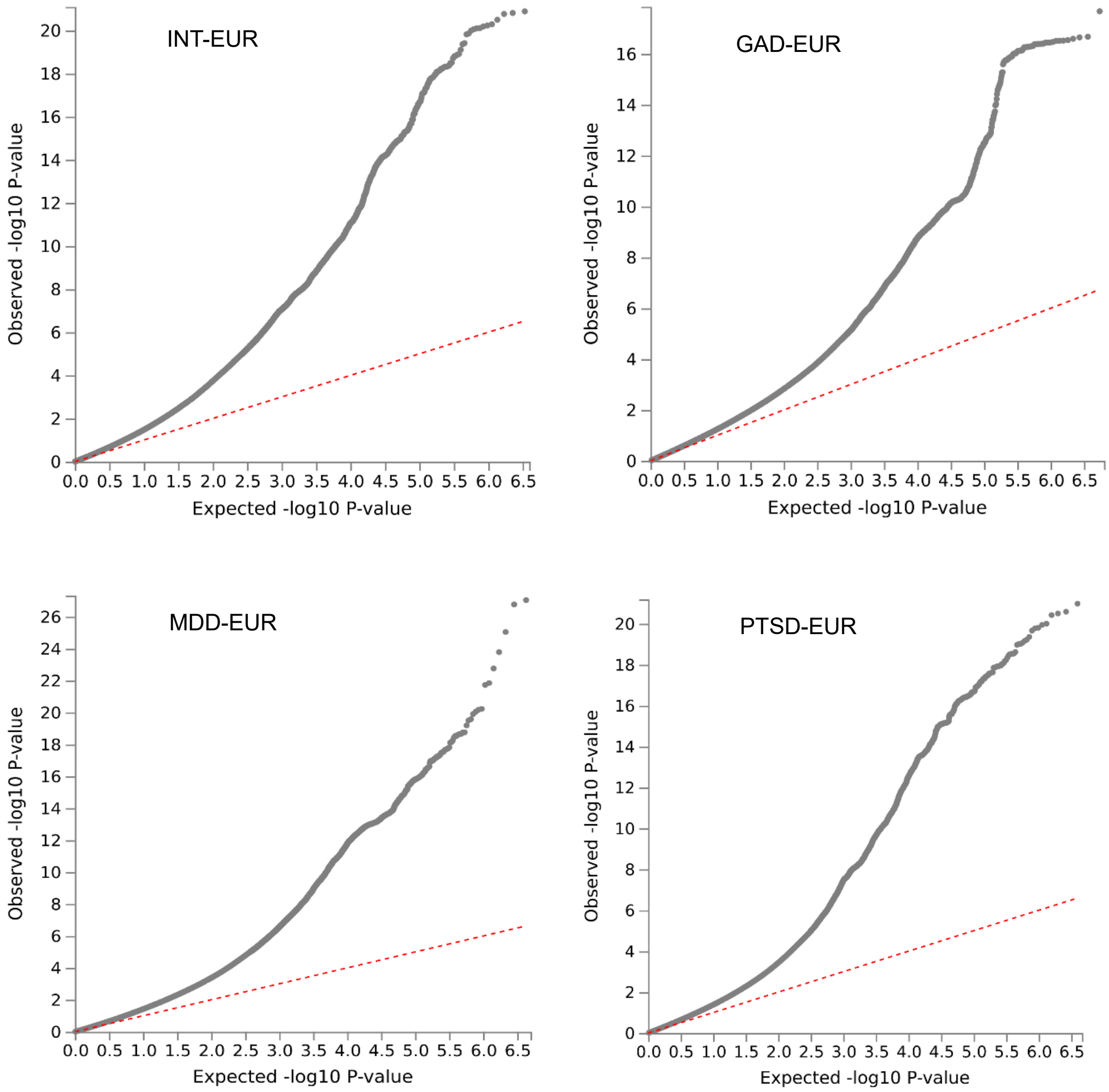


Figure S1. Manhattan plots for disorder-specific internalizing phenotypes and QQ plots of GWAS for shared internalizing disorder factor and disorder-specific internalizing phenotypes in European ancestry. INT: internalizing disorder factor; GAD: Generalized anxiety disorder; MDD: Major depressive disorder; PTSD: posttraumatic stress disorder; EUR: European ancestry.

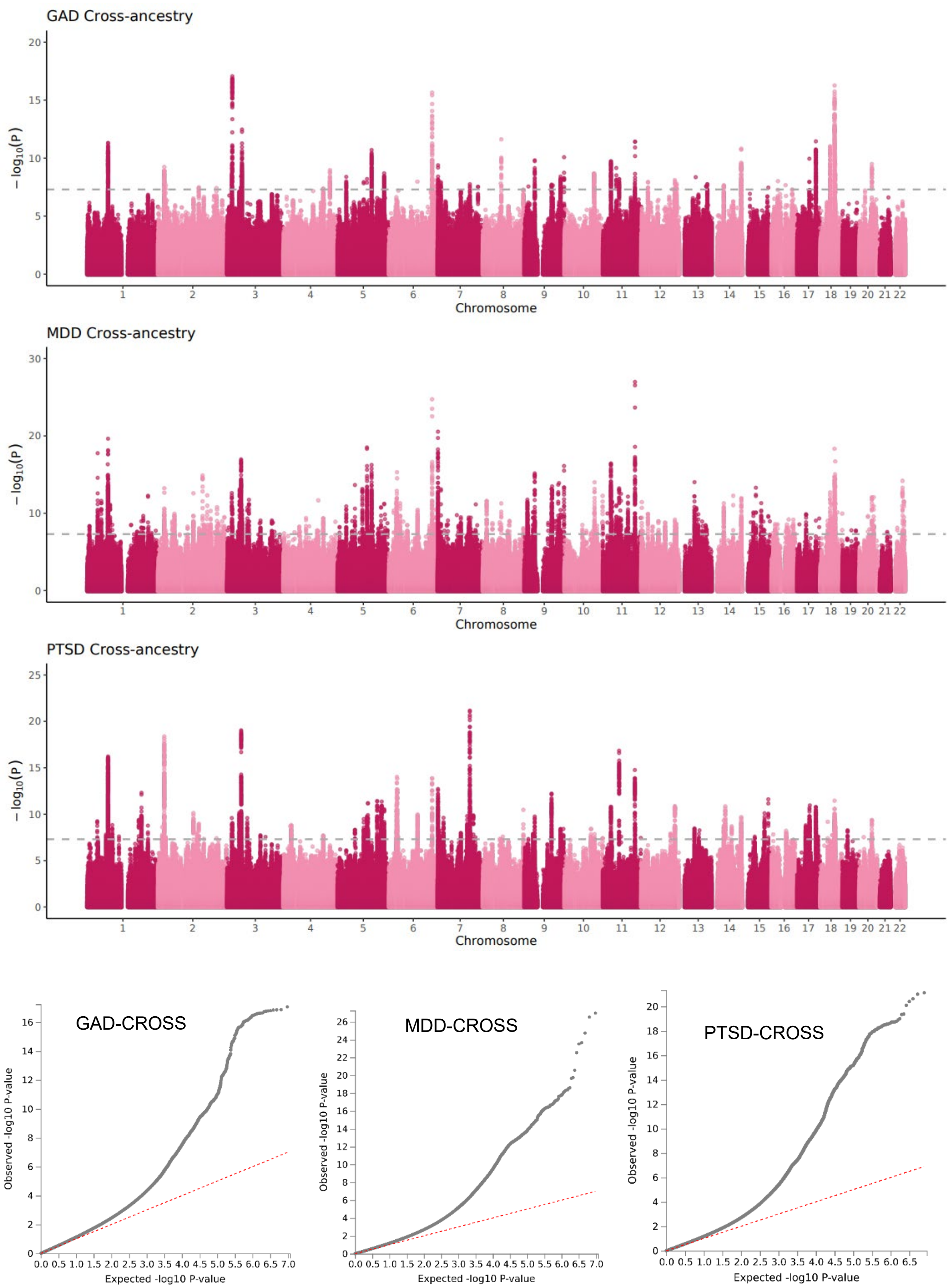
GAD Cross-ancestry
MDD Cross-ancestry
PTSD Cross-ancestry
− log10(P)
Chromosome
1 2 3 4 5 6 7 8 9 10 11 12 13 14 15 16 17 18 19 20 21 22
GAD-CROSS
MDD-CROSS
PTSD-CROSS
Observed -log10 P-value
Expected -log10 P-value

Figure S2. Manhattan plots and QQ plots of GWAS for internalizing phenotypes. Manhattan plots were based on GWAS effects in cross-ancestry meta-analysis. QQ plots were based on GWAS in cross ancestry. GAD: Generalized anxiety disorder; MDD: Major depressive disorder; PTSD: posttraumatic stress disorder.

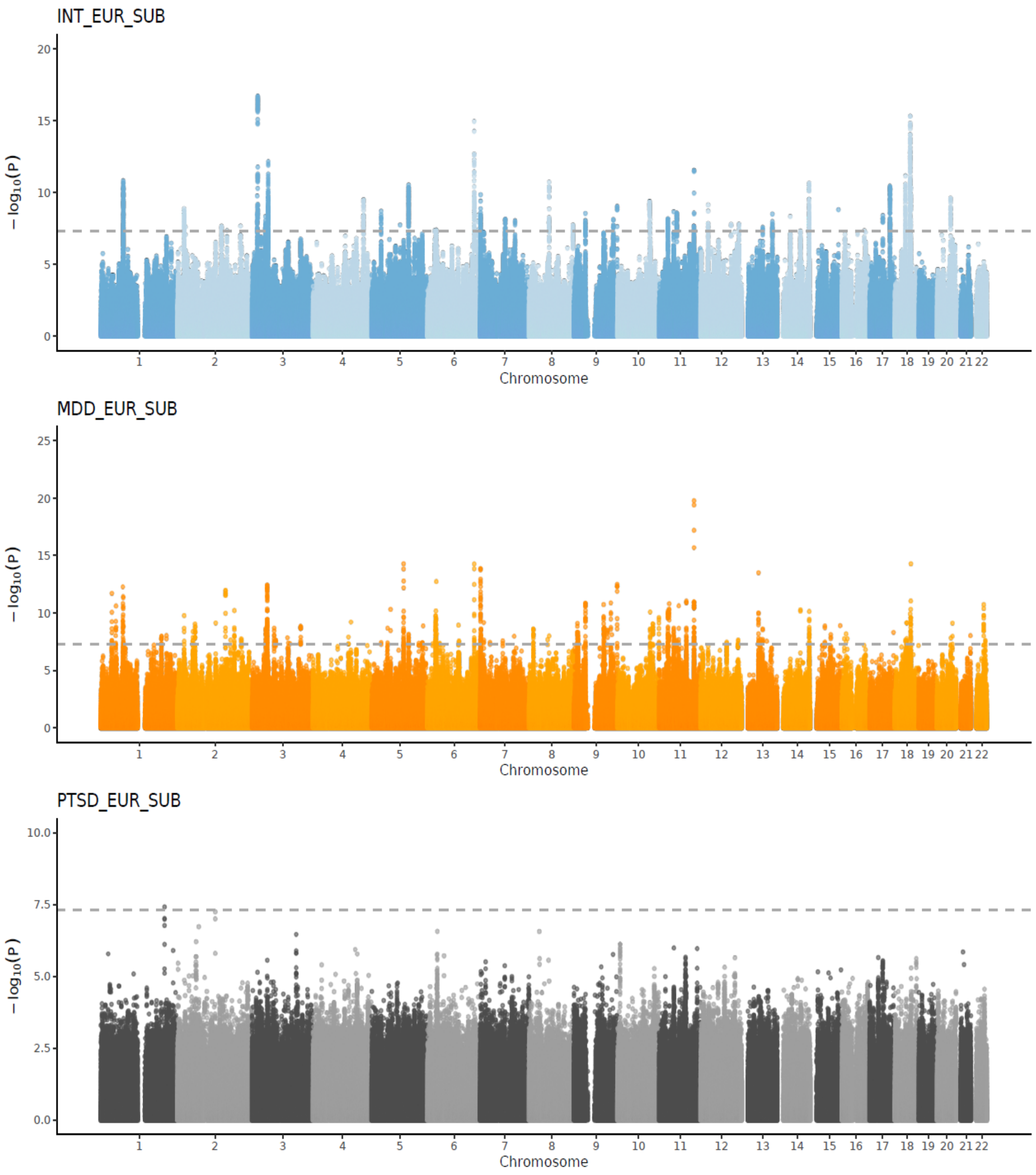

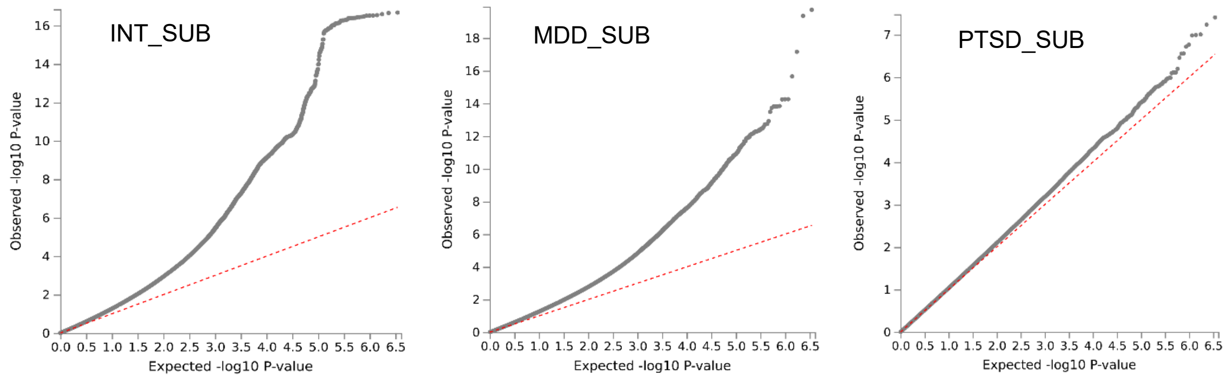


Figure S3. Manhattan plots and QQ plots of GWAS by subtraction for internalizing phenotypes. Manhattan plots were based on GWAS effects in GWAS by subtraction analysis. QQ plots were based on GWAS by subtraction in European ancestry. INT: internalizing disorder factor; MDD: Major depressive disorder; PTSD: posttraumatic stress disorder; EUR: European ancestry; SUB: subtraction.

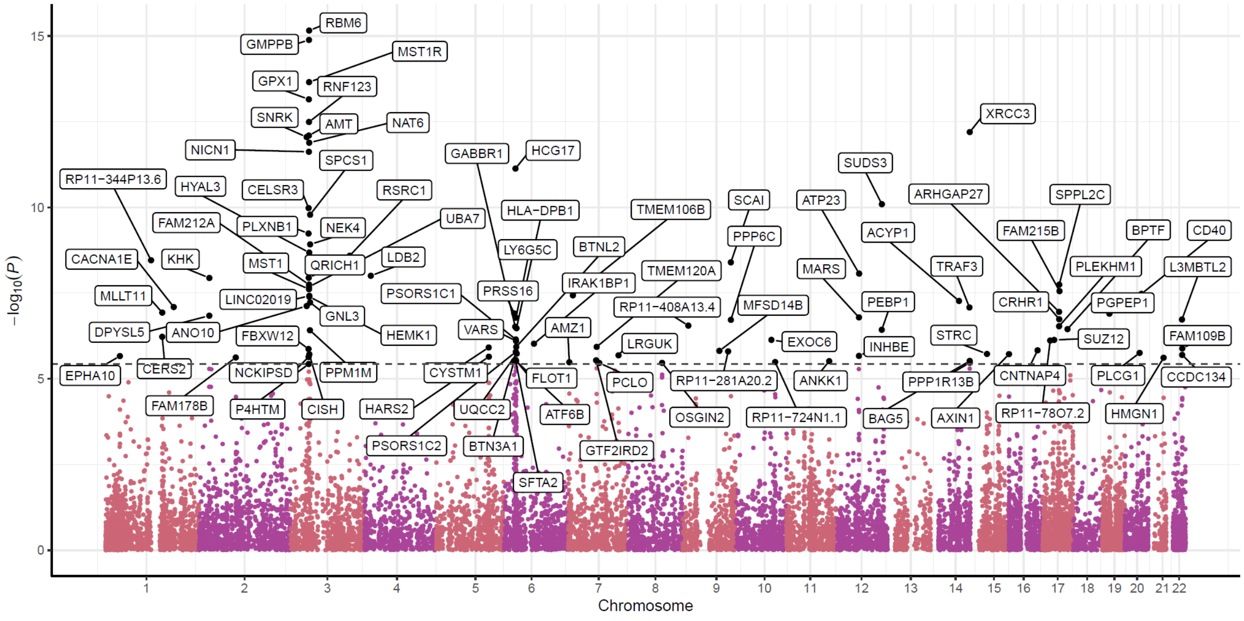


Figure S4. Manhattan plot of TWAS for internalizing disorder factor. Point plot was based on multi-tissue analysis using 13 brain tissues.

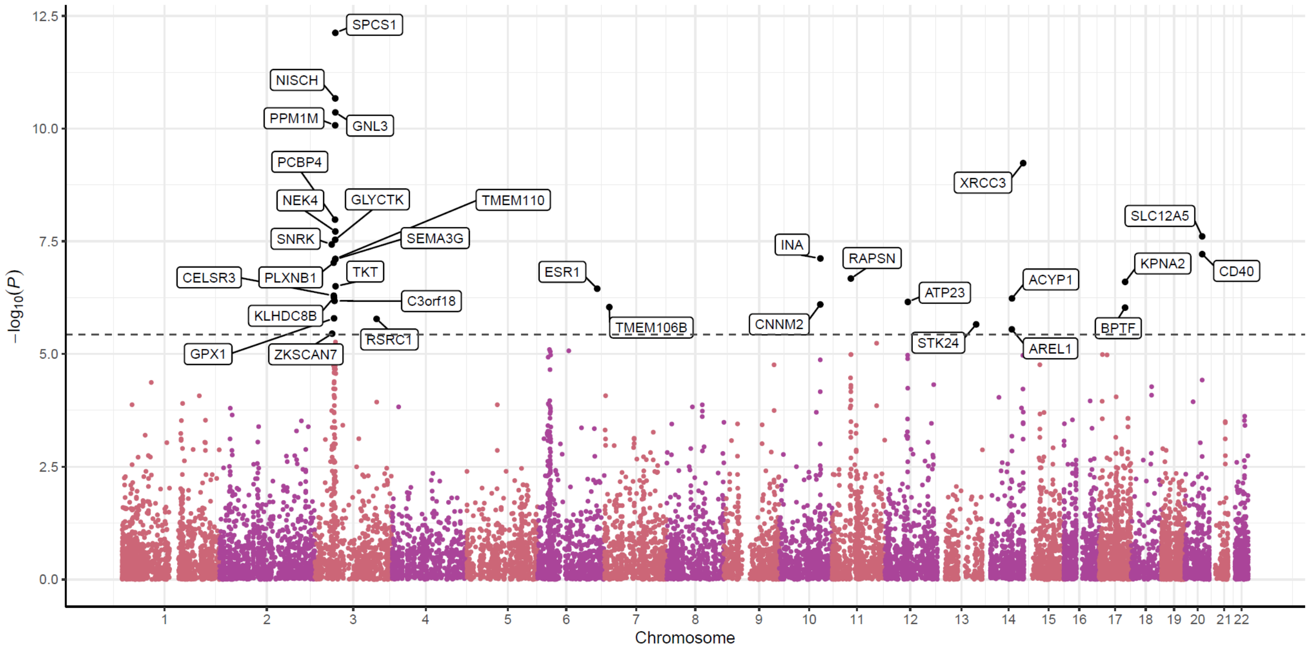


Figure S5. Manhattan plot of TWAS for generalized anxiety disorder in European desent. Point plot was based on multi-tissue analysis using 13 brain tissues.

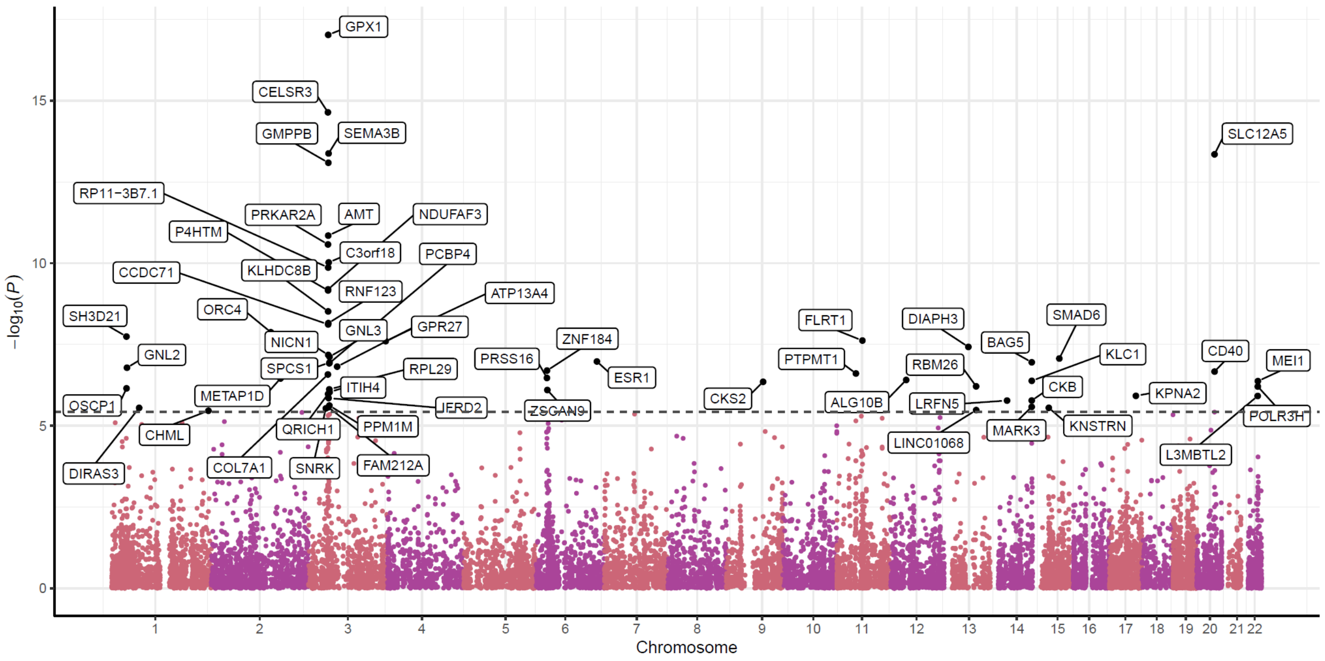


Figure S6. Manhattan plot of TWAS for major depressive disorder in European desent. Point plot was based on multi-tissue analysis using 13 brain tissues.

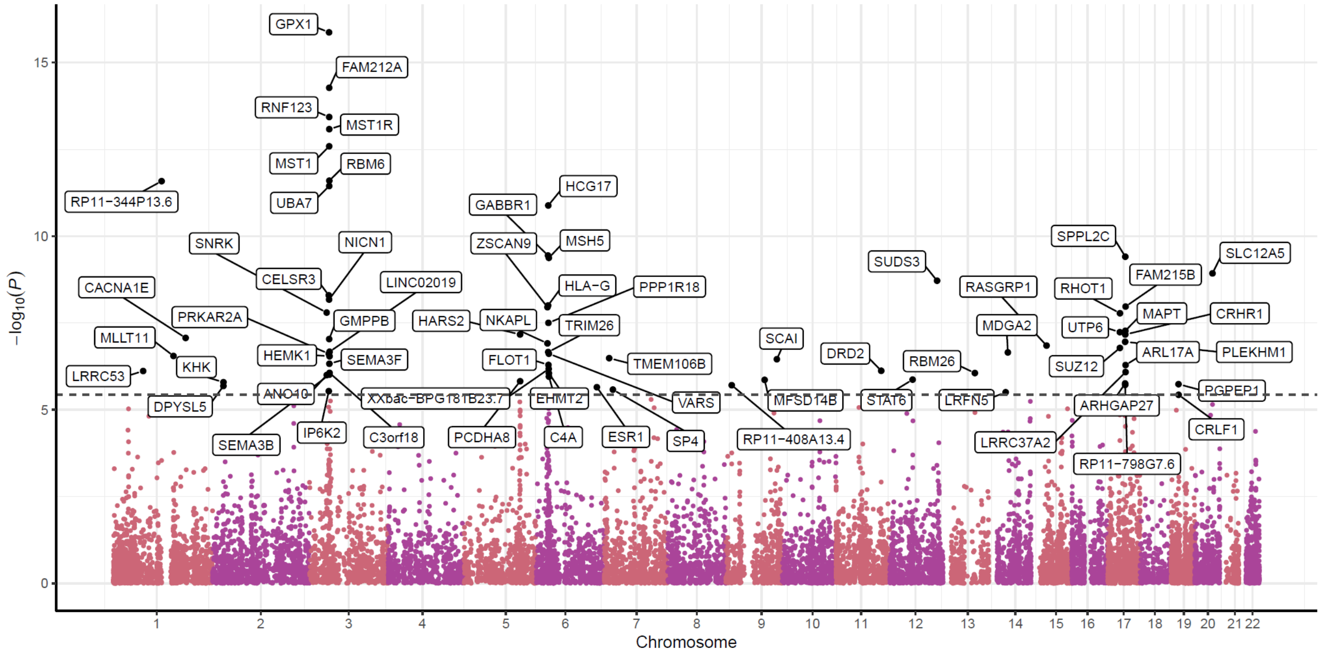


Figure S7. Manhattan plot of TWAS for posttraumatic stress disorder in European desent. Point plot was based on multi-tissue analysis using 13 brain tissues.

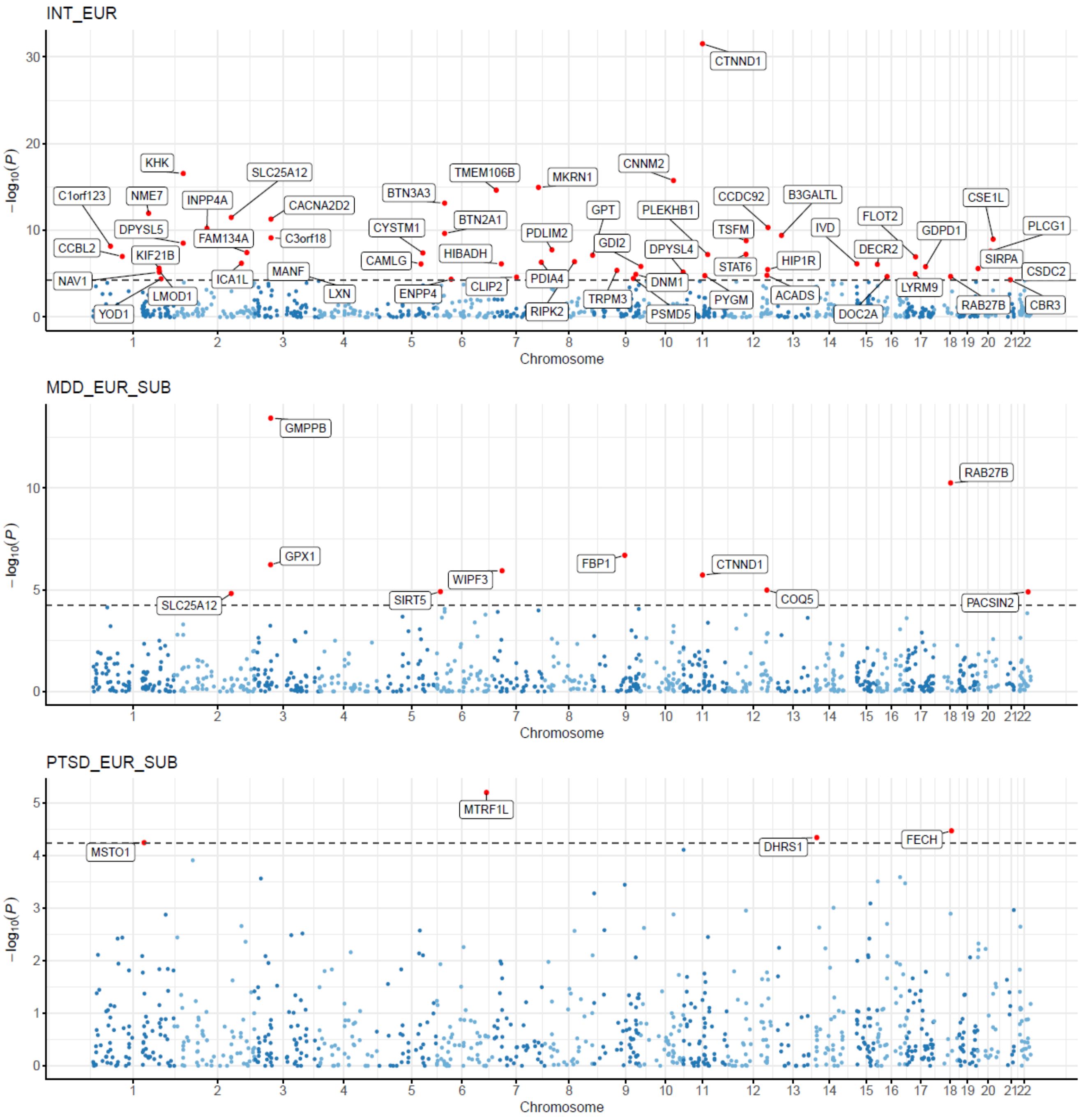


Figure S8. Manhattan plots of PWAS for internalizing phenotypes in European desent. Significant proteins associated with internalizing phenotypes were labeled. INT: internalizing disorder factor; MDD: Major depressive disorder; PTSD: posttraumatic stress disorder; EUR: European ancestry; SUB: subtraction.

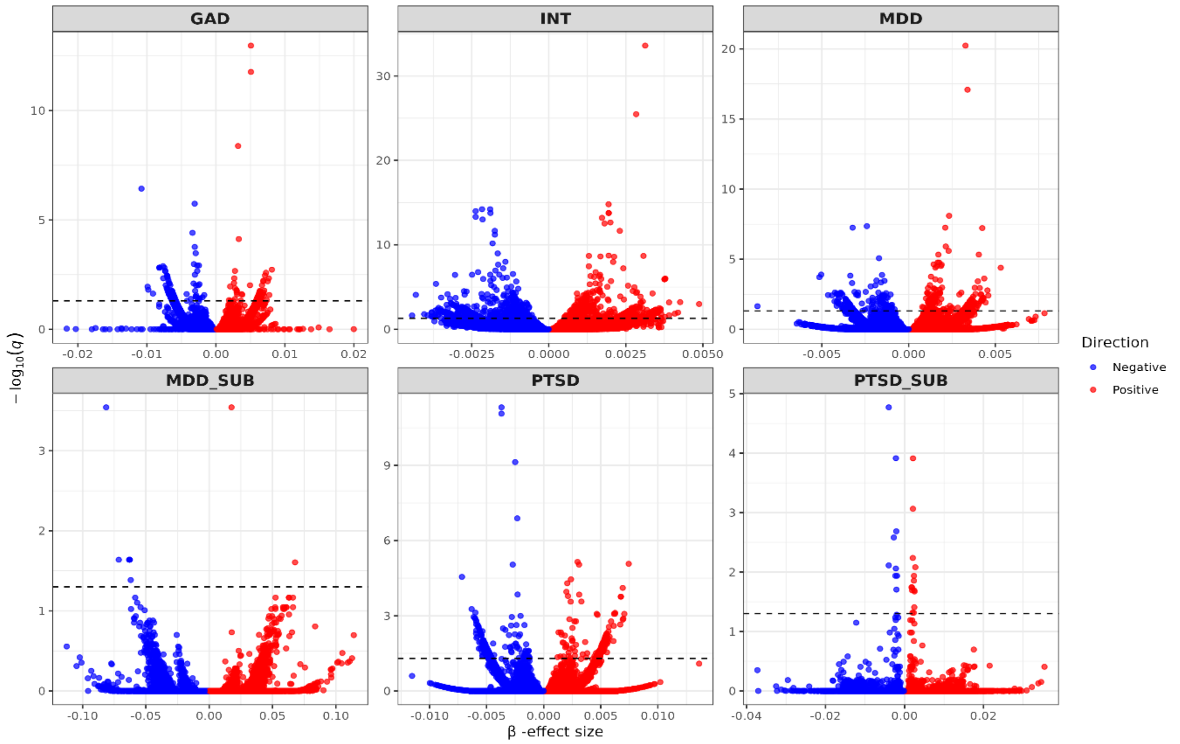


Figure S9. SMR results for internalizing phenotypes in individuals of European ancestry based on cross-tissue (brain–blood) meta-analysis after FDR correction. INT: internalizing disorder factor; GAD: Generalized anxiety disorder; MDD: Major depressive disorder; PTSD: posttraumatic stress disorder; EUR: European ancestry; SUB: subtraction.

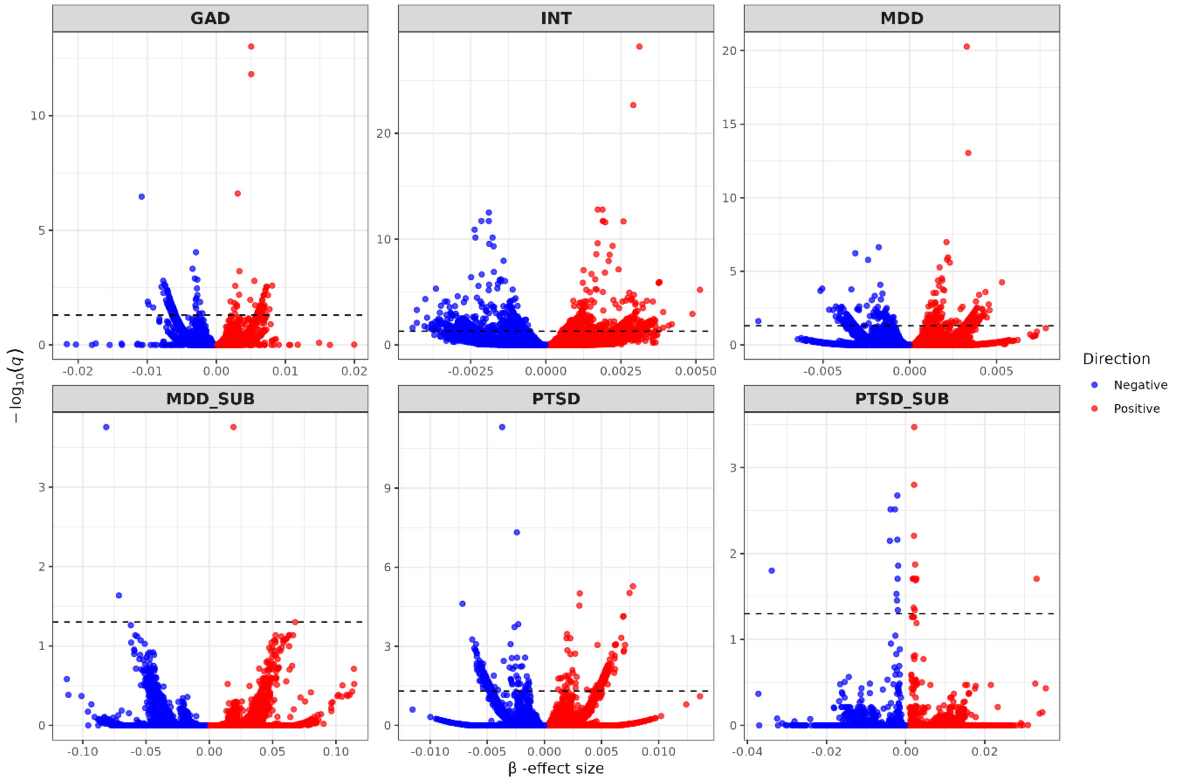

Figure S10. SMR results for internalizing phenotypes in individuals of European ancestry based on cross- blood tissue meta-analysis after FDR correction. INT: internalizing disorder factor; GAD: Generalized anxiety disorder; MDD: Major depressive disorder; PTSD: posttraumatic stress disorder; EUR: European ancestry; SUB: subtraction.

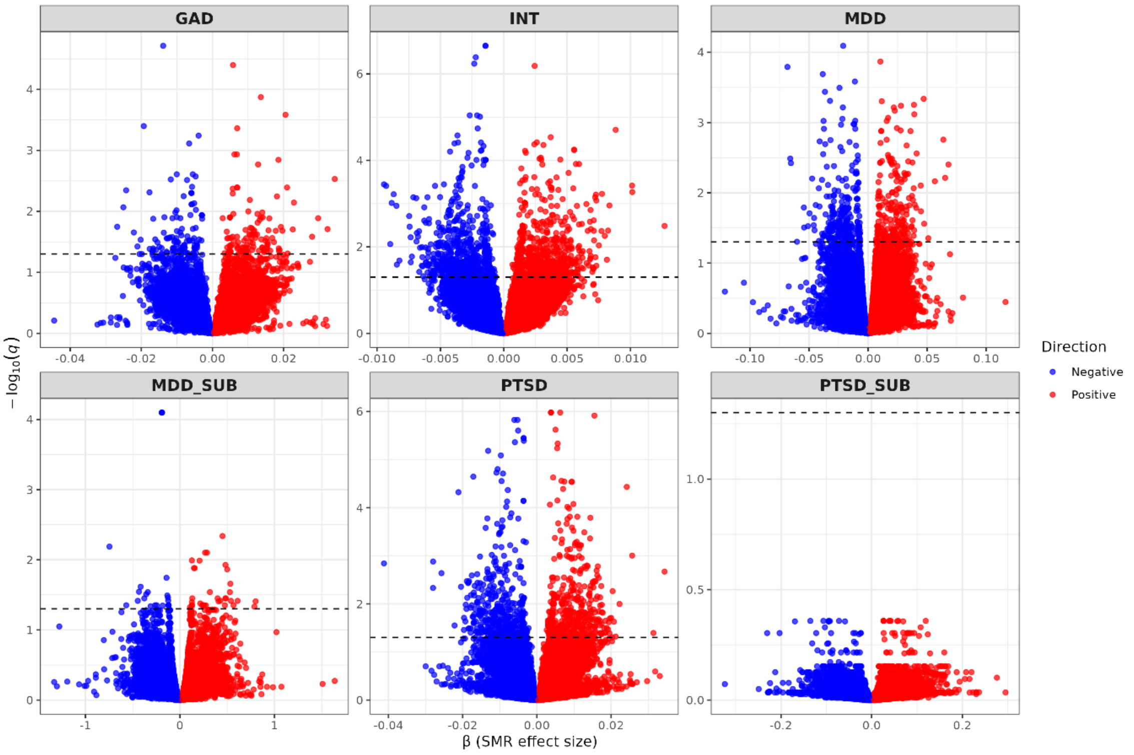


Figure S11. SMR results for internalizing phenotypes based on brain tissue meta-analysis in European ancestry after FDR correction. INT: internalizing disorder factor; GAD: Generalized anxiety disorder; MDD: Major depressive disorder; PTSD: posttraumatic stress disorder; EUR: European ancestry; SUB: subtraction.

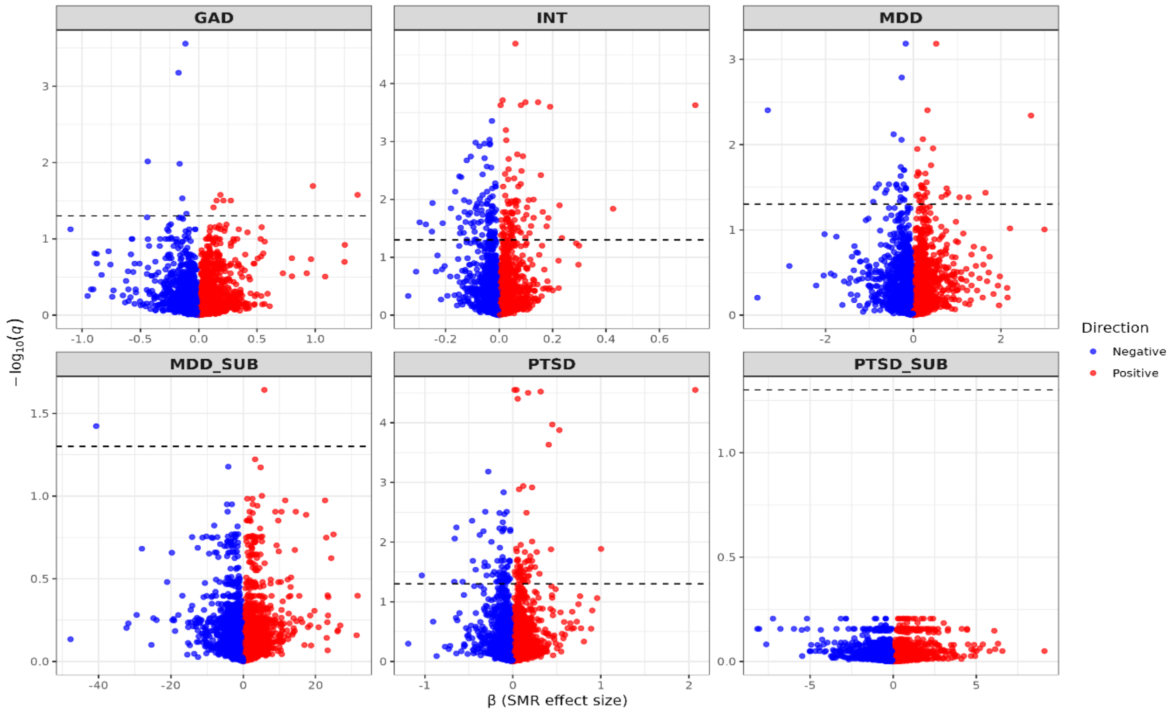


Figure S12. SMR results for internalizing phenotypes based on fetal brain tissue meta-analysis in European ancestry after FDR correction. INT: internalizing disorder factor; GAD: Generalized anxiety disorder; MDD: Major depressive disorder; PTSD: posttraumatic stress disorder; EUR: European ancestry; SUB: subtraction.

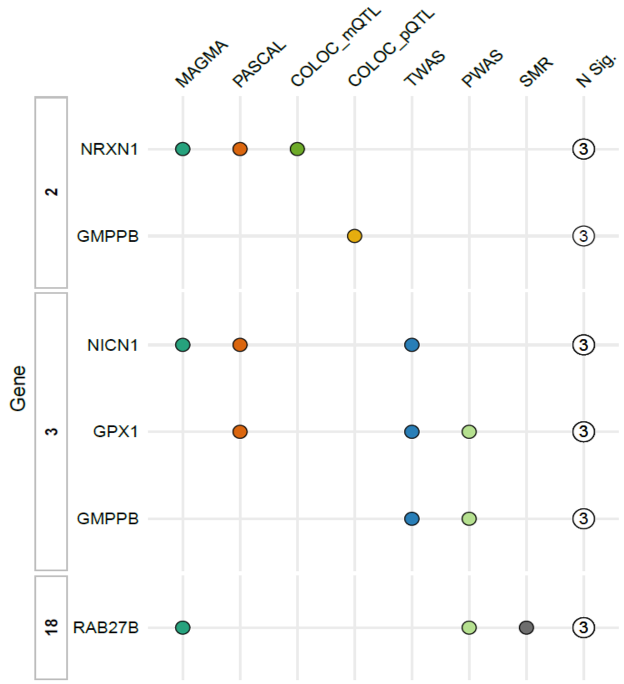


Figure S13. Top genes associated with major depressive disorder GWAS by subtraction in European desent across ten complementary approaches. COLOC_mQTL: Colocalization analysis with methylation quantitative trait loci; COLOC_pQTL: Colocalization analysis with brain protein quantitative trait loci; MAGMA: Multi-marker Analysis of GenoMic Annotation; PASCAL: Pathway

Scoring Algorithm; PWAS: Proteome-Wide Association Study; SMR: Summary-based Mendelian randomization; TWAS: Transcriptome-Wide Association Study.

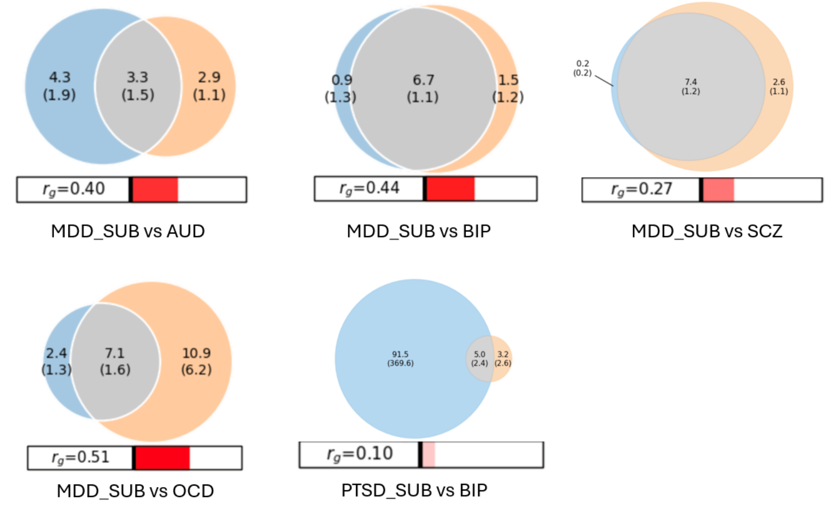


Figure S14. Causal variants shared between major depressive disorder GWAS by subtraction and psychiatric disorders estimated by bivariate MiXeR model. AUD: Alcohol use disorder; BIP: bipolar disorder; MDD: Major depressive disorder; OCD: Obsessive-compulsive disorder; PTSD: posttraumatic stress disorder; SCZ: schizophrenia; SUB: subtraction.
